# Intraplaque haemorrhage quantification and molecular characterisation using attention-based multiple instance learning

**DOI:** 10.1101/2025.03.04.25323316

**Authors:** Francesco Cisternino, Yipei Song, Tim S. Peters, Maria Murach, Patrick Hart, Jose Verdezoto Mosquera, Laura Mäkitie, Mikko I. Mäyränpää, Luka Zivkovic, Roya Batool, Julian Louma, Abdalla Tarek Marei, Nikolaos Tsilimparis, Ana Karina de Oliveira, Roderick Westerman, Gert J. de Borst, Ernest Diez Benavente, Noortje A.M. van den Dungen, Petra Homoed-van der Kraak, Dominique P.V. de Kleijn, Joost Mekke, Michal Mokry, Gerard Pasterkamp, Hester M. den Ruijter, Evelyn Velema, Petra Ijäs, Marios K. Georgakis, Clint L. Miller, Craig A. Glastonbury, Sander W. van der Laan

**Author notes:** **Corresponding author(s):** *Dr. Sander W. van der Laan*, Central Diagnostic Laboratory, Division Biomedical genetics, Laboratories, and Pharmacy University Medical Center Utrecht, Utrecht University, Utrecht, The Netherlands Department of Genome Sciences, University of Virginia, Charlottesville, VA, USA., *Dr. Craig A. Glastonbury*, Genomics Research Centre, Human Technopole, Italy, Nuffield Department of Medicine, University of Oxford, Oxford, UK, *Dr. Clint L. Miller*, Department of Genome Sciences, Department of Public Health Sciences, Department of Biochemistry and Molecular Genetics, University of Virginia, Charlottesville, VA, USA. these authors contributed equally (Francesco Cisternino, Yipei Song, Tim S. Peters, Clint L. Miller, Craig A. Glastonbury, Sander W. van der Laan).

## Abstract

Intraplaque haemorrhage (IPH) destabilises atherosclerotic plaques and is associated with myocardial infarction and stroke. However, its identification and quantification remain limited by manual histological scoring. We introduce PHENOMICL, an attention-based additive multiple instance learning (MIL) framework trained on 2,595 carotid plaques to detect, localise, and quantify IPH across nine stains. Haematoxylin and Eosin (H&E), a routinely available stain, achieved strong single-stain performance (AUROC=0.86), while multi-stain ensemble models combining H&E+CD68 or Verhoeff-Van Gieson improved discrimination (AUROC=0.92). The outputs enabled slide-level classification and generated spatially resolved patch-level probability maps providing continuous estimations of IPH area. Model-IPH outperformed manual scoring for classifying preoperative symptoms and major adverse cardiovascular events (MACE). Integration with bulk, single-cell, and spatial transcriptomics linked IPH to inflammatory, foam-cell and extracellular matrix remodelling, including TNF-alpha signalling. Cell-cell communication identified the CCL-ACKR1 axis linking macrophage activity to angiogenesis and IPH. PHENOMICL establishes a scalable, interpretable framework converting routine histology images into quantitative phenotypes.

## Introduction

Atherosclerosis is a chronic inflammatory process affecting arterial walls, leading to lipid, inflammatory cells, and fibrous element accumulation to atherosclerotic plaques^1,2^. These plaques are heterogeneous, with a multitude of endophenotypes present, such as lipid-rich necrotic cores, fibrous caps, calcification, and intraplaque haemorrhage (IPH), each offering unique insights into plaque stability and the likelihood of major adverse cardiovascular events (MACE)^3,4^.

IPH marks the transition of a stable plaque into a vulnerable one, prone to rupture, and often associated with thrombus formation and MACE^5,6^. The process of IPH is multifaceted, involving the formation of leaky neo-vessels due to the lack of smooth muscle cells and endothelial gap junctions^7^. In the hypoxic environment of the plaque, IPH stimulates neutrophils to secrete angiogenic factors and lipid peroxidation products, while macrophages scavenge leaked erythrocytes, releasing haemoglobin and iron^8^. These processes underscore IPH’s complexity and critical role in plaque vulnerability (**Figure 1A**).

**Figure 1:**
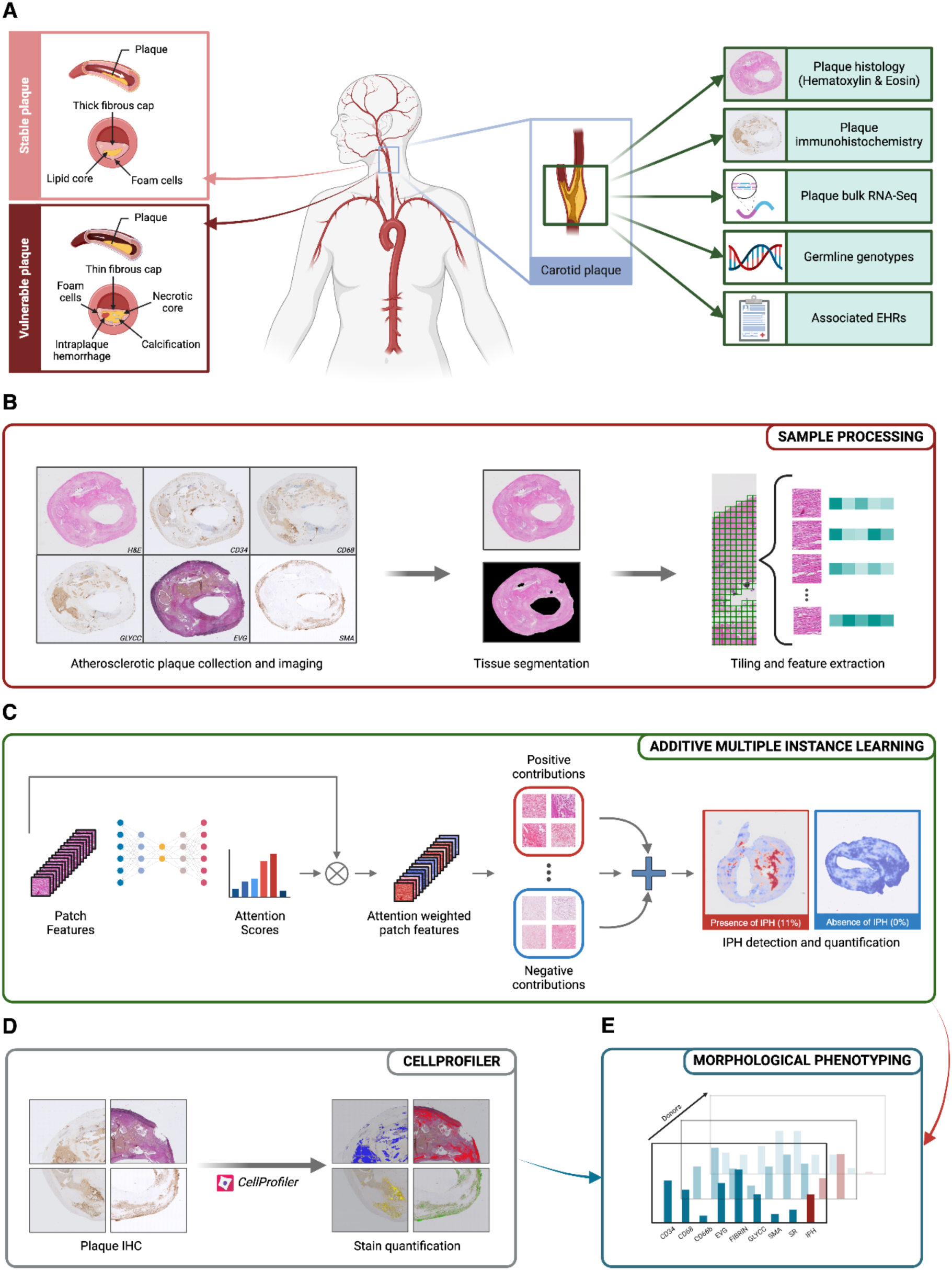
Schematic research overview. (**A**) Carotid plaque vulnerability shows up with specific biomarkers, like development of intraplaque haemorrhage (IPH) and a necrotic core (Left). In this study carotid plaques were collected during endarterectomy. Samples were stained with Hematoxylin & Eosin and 8 additional histological and immuno-histochemical (IHC) markers, plaque bulk RNA-Seq were collected, together with germline genotypes; electronic health records are available for the whole cohort (Right). (**B**) After scanning the stained tissue samples, whole-slide images (WSI) were pre-processed by segmenting tissue regions from background, patching and performing feature extraction. (**C**) Attention-based additive multiple instance learning was employed to classify, quantify and localize IPH in the tissue; coupling this image-derived biomarker with stain quantification from CellProfiler^9,10^ (**D**) resulted in a complete morphological phenotyping (**E**) of carotid plaques.

To assess IPH and other plaque phenotypes, histopathological examinations rely on various stains, each tailored to reveal specific cellular or extracellular components^10^. These include: Hematoxylin and Eosin (H&E) providing general plaque morphology; Elastic van Gieson (Verhoeff-van Gieson, EVG) and Picrosirius Red (SR) highlighting elastic and collagen fibres, critical for assessing structural remodelling in response to IPH^11^ CD34 and Smooth Muscle Actin (SMA) to assess neovascularization and fibrous cap integrity^12 13^; CD68 and CD66b to identify macrophage and neutrophil infiltration^14,15^; and glycophorin C (GLYCC) and FIBRIN to specifically localize erythrocytes and blood clots^10^. While manual scoring of IPH (manual-IPH) uses Hematoxylin and Eosin, combining stains can provide a more comprehensive picture of the cellular and structural IPH landscape within atherosclerotic plaques. However, to date, no systematic comparison has been made to assess the utility or redundancy of these stains in capturing IPH.

Phenotyping plaques using individual stains is labour intensive, subjective, and prone to high inter-sample variability, necessitating the development of advanced computational techniques to enhance phenotyping. Machine learning (ML) has emerged as a powerful tool for analyzing complex biomedical images, offering objective, high-throughput, and quantitative phenotyping of atherosclerotic plaques^16–19^. Machine learning models have been successfully employed for automated segmentation of plaque components such as lipid cores, fibrous caps, and calcifications in various imaging modalities^20–23^. ML has been applied to analyze inflammatory cell infiltration in tissue samples^24,25^ and integrate imaging data with genomics and proteomics to identify molecular pathways associated with several types of disease^26–28^.

We present an attention-based additive multiple instance learning (MIL) model, PHENOMICL, to automate, spatially localize and quantify IPH (model-IPH) in 2,595 plaques using 9 distinct histological stains (**Figure 1B,C**). Quantifying model-IPH area allows us to characterize whether IPH abundance relates to symptom severity and secondary outcomes. Besides IPH quantification, we systematically characterise the composition of all histological stains to capture the full diversity of cellular and extracellular plaque components (**Figure 1D**), enabling a more comprehensive and automated phenotyping of plaques (**Figure 1E**). Notably, H&E histology alone provides excellent classification performance, suggesting that PHENOMICL can be used with the most widely available staining protocol. Rigorous validation against traditional histopathology demonstrates machine learning’s potential to improve accuracy, efficiency, and reproducibility in atherosclerosis phenotyping. Additionally, PHENOMICL was validated across other vascular beds, independent cohorts, and different methodological annotations with spatially marked IPH and microhaemorrhage regions. Further characterization of molecular properties of plaques with IPH was performed through differential gene expression analysis, highlighting macrophage-related pathways, TNF-α signaling, estrogen suppression, and the role of *HMOX1* in foam cells, which was validated by single-cell sequencing and follow-up IHC staining. Spatial transcriptomic analyses confirmed the localization of model-IPH-associated gene signatures, and cell type deconvolutions revealed that macrophage populations play an important role within the regions of haemorrhage. Cell- cell communication analysis further identified the CCL-ACKR1 axis, signaling from macrophages to ACKR1-expressing endothelial cells, as a potential mediator linking macrophage activity to angiogenesis and IPH. This study presents a robust method for quantifying IPH plaque pathology and composition and investigating its molecular and cellular heterogeneity.

## Results

### Cohort and data description

This study utilizes the Athero-Express Biobank, an ongoing study conducted in two Dutch tertiary hospitals with the objective to investigate the etiological value of plaque characteristics for long-term clinical outcomes^29^. Carotid plaques were obtained from 2,595 patients (1,803 males, 792 females) undergoing carotid endarterectomy (CEA). There was a noticeable difference between males and females in baseline clinical characteristics and plaque morphology, including significantly lower prevalence of manual-IPH in females compared to males (53.1% vs 64.9% respectively). A comprehensive breakdown of the cohort is provided in **Supplemental Table 1**. During the 3-year follow-up period, 452 (25.6%) males and 171 (22.1%) females reached a composite endpoint (see Methods).

### Automated detection and quantification of Intraplaque haemorrhage from nine histological stains

Intraplaque haemorrhage is typically detected by identifying the presence of red blood cells (RBC), either through H&E staining or through specific markers of erythrocytes (e.g. glycophorin C). However, the manual detection and quantification of IPH in samples can be hard to achieve at scale. This motivated the development of an automated method to quantify IPH in carotid plaque histology. Nine stain-specific additive MIL models were trained using the manual-IPH label as ground truth (**Supplemental Figure 1**). Train, validation, and test set sizes alongside AUROC (area under the receiver operating characteristic curve), accuracy, and F1-score metrics after 10-fold cross-validation are reported in **Figure 2** and **Supplemental Table 2.** H&E achieved the highest single stain performance (AUROC = 0.86 ± 0.017), followed by FIBRIN (AUROC = 0.85 ± 0.035) and EVG (AUROC = 0.84 ± 0.031). Importantly, whilst EVG had a comparable number of patients compared to H&E (n=1,743 versus n=1,715, respectively), there were 2.7x more H&E whole slide images as compared to FIBRIN (n=628), suggesting FIBRIN is a highly specific stain to measure IPH accurately. To assess whether any gain in performance can be achieved by leveraging multiple stains, we fit logistic regression ensemble models combining the probability of having model-IPH across multiple stains. For each logistic regression ensemble model, we combined the probabilities from H&E (the highest-scoring single stain model) with those from another stain. Train and test set sizes alongside AUROC, accuracy, and F1-score metrics after 10-fold cross-validation are reported in **Figure 2** and **Supplemental Table 3.** The combinations achieving the best AUROC were H&E+CD68 (AUROC = 0.92 ± 0.016) and H&E+EVG (AUROC = 0.92 ± 0.017), emphasising the importance of both macrophages and vascular remodelling in IPH.

**Figure 2:**
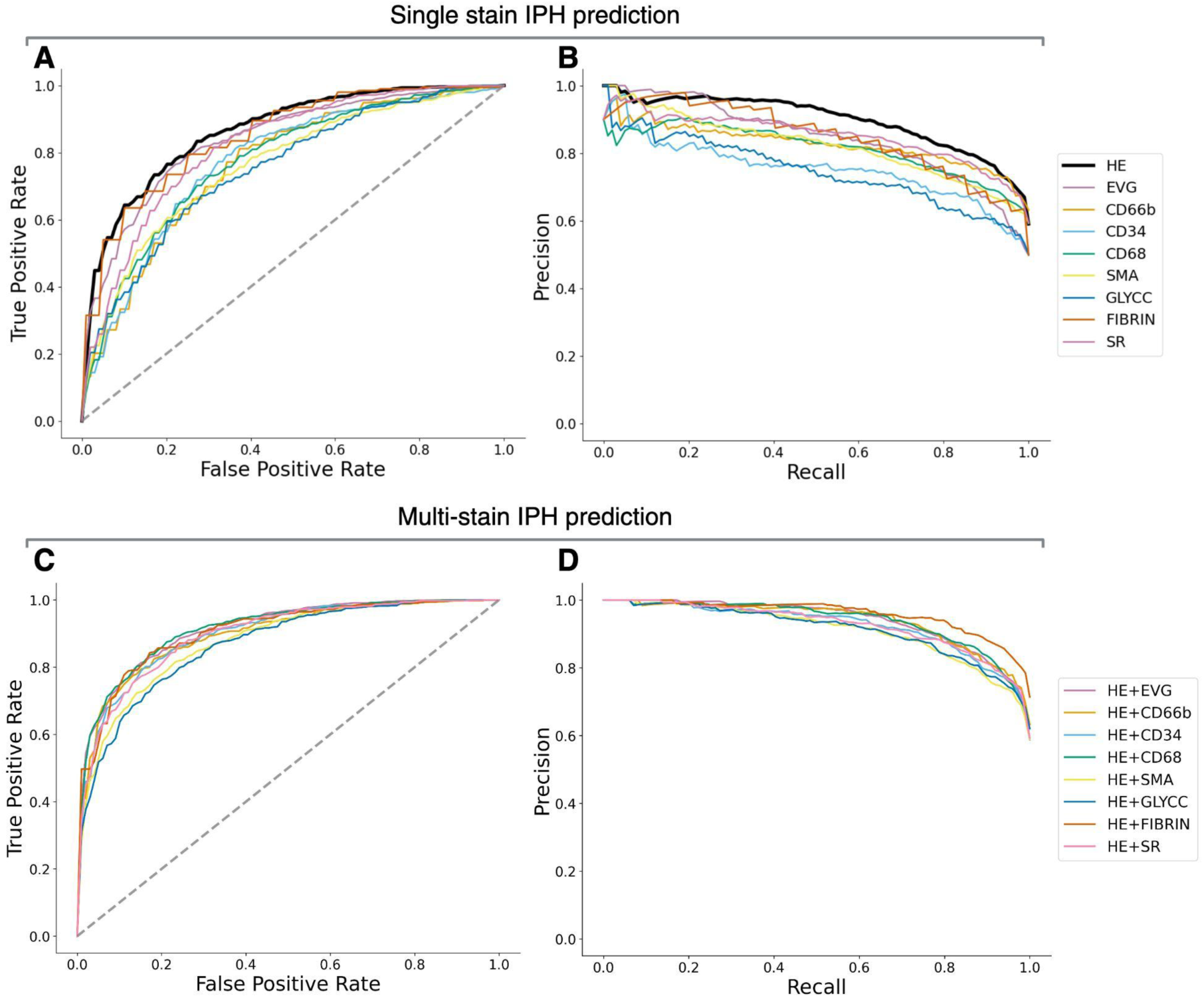
Additive MIL model performance. Receiver Operating Characteristic (ROC) curves (**A, C**) and Precision-Recall (PR) curves (**B, D**) for classification models trained using single histological stains (**A, B**) and multi-stain logistic regression ensemble models combining Hematoxylin & Eosin (H&E) with each of the other stains (**C, D**). The top panels show performance from models trained on individual stains: H&E, Elastin Verhoeff–Van Gieson (EVG), CD66b, CD34, CD68, Smooth Muscle Actin (SMA), glycophorin C (GLYCC), Fibrin, and Sirius Red (SR). The bottom panels present results from logistic regression ensemble combinations: H&E+EVG, H&E+CD66b, H&E+CD34, H&E+CD68, H&E+SMA, H&E+GLYCC, H&E+Fibrin, and H&E+SR. The diagonal dashed line in ROC curves represents the performance of a random classifier. H&E represent the best stain for classifying the presence of IPH. For the multi-stain comparison, H&E+CD68 achieved the highest AUROC score, while H&E+FIBRIN obtained the highest F1-score.

Our additive MIL framework allows us to leverage instance patch-level contributions to calculate the overall probability for the sample (model-IPH). These patch contributions, which may be positive, negative, or irrelevant to the final outcome, provided clear interpretability of the precise morphological features associated with IPH. This approach allows both the detection of IPH and its localization and quantification in terms of area of the plaque occupied by IPH (derived IPH area), even when the ground truth label is binary or categorical. These localized IPH areas show good concordance with GLYCC staining (**Figure 3**) and manual annotations (**Supplemental Figure 3**).

**Figure 3:**
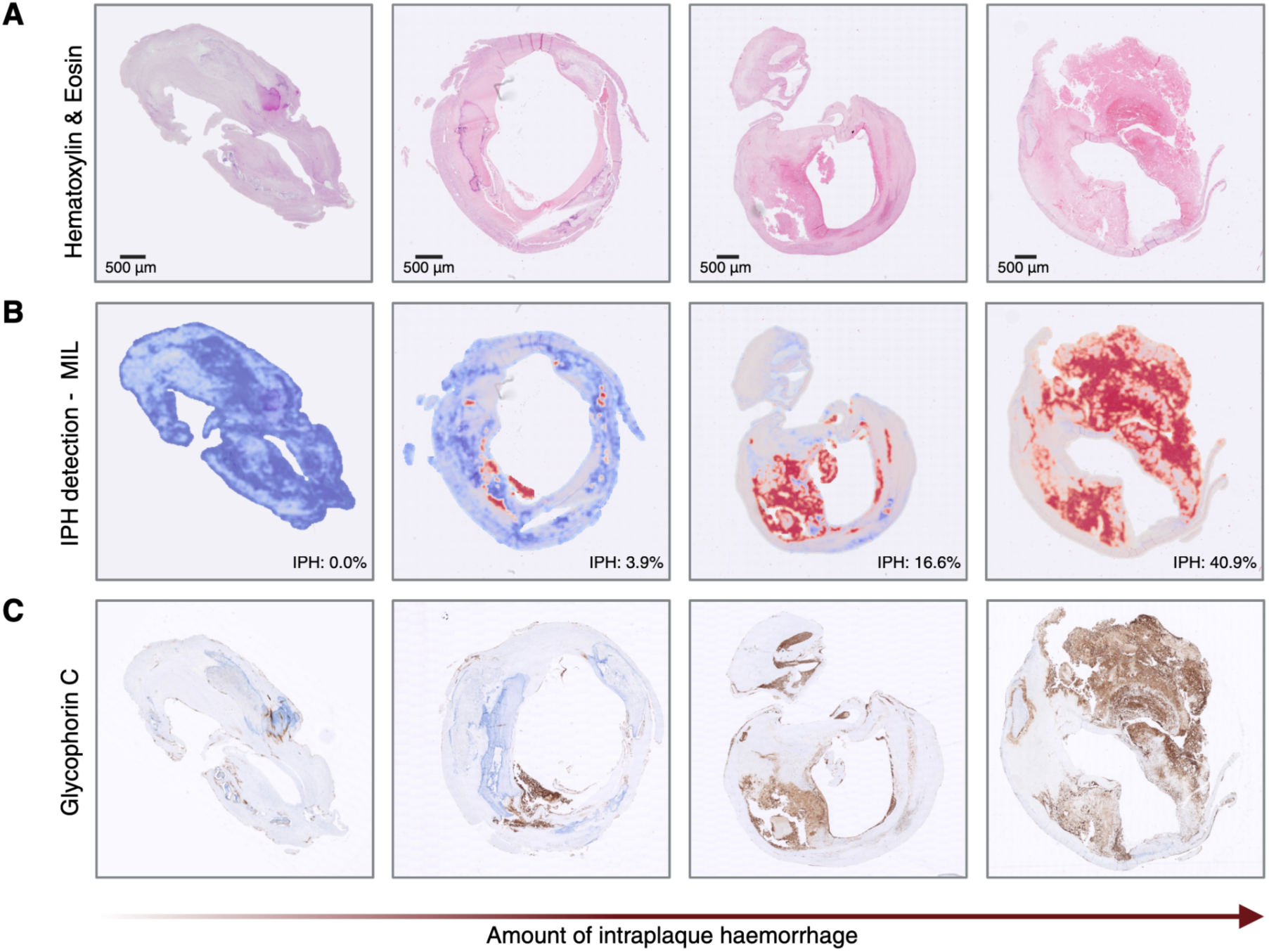
Additive MIL attention heatmaps. Intraplaque haemorrhage localization and quantification, expressed as a percentage of total plaque area (**B**) from H&E images (**A**) compared to glycophorin C staining (**C**).

The single stain H&E model was used to derive the IPH area across all 2,220 plaque images. The percentage of plaque area occupied by regions of IPH varied substantially (n=2,220, mean = 10%, SD = 1%, range: 0–63.6%) between subjects. The extent of IPH area is, on average, higher and more variable in men (mean = 11%, SD = 10%) than in women (mean = 7%, SD = 9%) (p-value = 6.64×10^−25^), suggesting IPH area is sexually dimorphic. As reported by several studies^30–32^, plaque rupture is more common in older women and men with high cholesterol levels, whereas younger women tend to develop plaque erosion. This trend is confirmed in our cohort, where younger (age < 60, n=125) women have a significantly lower mean IPH area than older (age ≥ 60, n=550) women (mean = 5% vs mean = 8%, p-value = 1.01×10^−5^). IPH is also more present in plaques with >10% of fat (mean = 14%, SD = 11%) than in plaques with a low fat percentage (mean = 10%, SD = 9%) (**Supplemental Figure 4)**. Finally, we find that IPH area significantly correlates positively with manually counted neutrophils (r = 0.41, p-value = 9.18×10^−13^) and negatively with manually counted smooth muscle cells (r = -0.29, p-value = 1.92×10^−31^). These findings collectively highlight the heterogeneity of atherosclerosis manifestations across age, sex, plaque fat content and cellularity.

### Validation of automated IPH detection across vascular beds, independent cohorts and manual scoring methods

To test generalizability, PHENOMICL was used to detect IPH across vascular beds and external cohorts, without retraining. The model maintained a solid classification performance on femoral endarterectomy H&E samples (AUROC = 0.813; **Supplemental Table 4; Supplemental Figure 2**) and the external HeCES2 carotid cohort (AUROC = 0.781; **Supplemental Table 5; Supplemental Figure 2**). Additional spatial localization validation against expert annotations in the AtherOMICS cohort demonstrated strong slide-level correlation between model-derived and manually annotated IPH areas (r = 0.89, p-value = 1.88×10^−49^). We looked at the alignment of AtherOMICS manual annotation with model-IPH heatmap across varying thresholds for the PHENOMICL model (the minimum patch-level score required to classify a patch as IPH-positive; **Supplemental Table 6**). At a threshold of 0.75, patch-level spatial agreement (DICE score) and sensitivity increased substantially with IPH area size (**Figure 4; Supplemental Table 7**). The correlation between DICE score and model-IPH area size (r = 0.655, p-value = 1.14×10^−17^) confirms that performance degradation in small lesions is systematic rather than random and is consistent with the patch resolution of the model. Meanwhile, high specificity across all lesion sizes (mean = 0.987 ± 0.013), confirms that the model rarely misclassifies non-IPH tissue as IPH positive.

**Figure 4:**
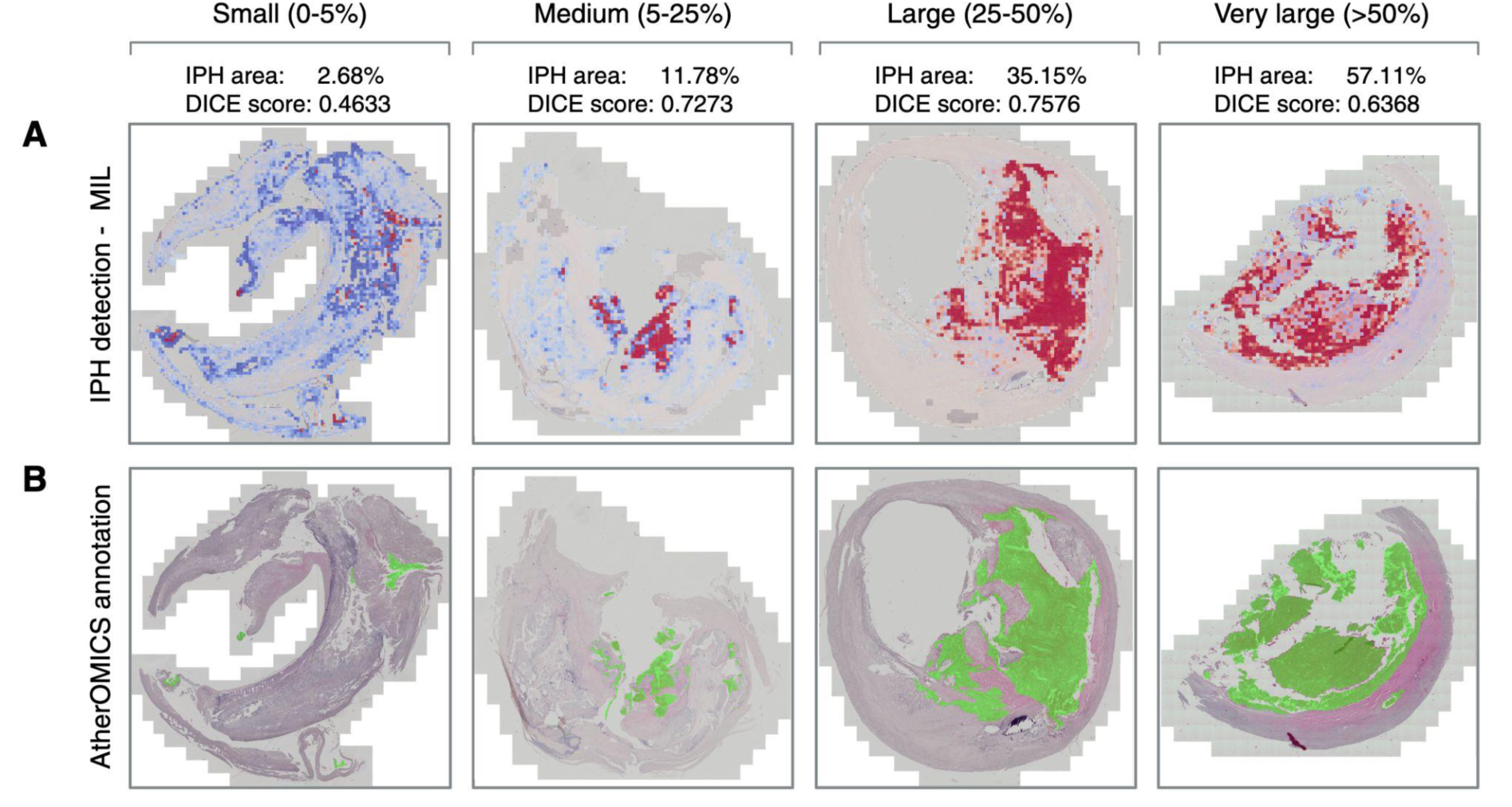
Additive MIL attention heatmaps validation using external AtherOMICS cohort. Intraplaque haemorrhage localization from H&E images (**A**) using the model compared to AtherOMICS manually annotated overlays (**B**), showing that the additive MIL model accurately localizes IPH regions based on H&E stained images.

Together, these external validation results demonstrate that PHENOMICL generalises well across vascular beds, independent cohorts, and scoring methodologies, with residual limitations in spatial agreement that could be explainable by lesion size and the proportionally higher prevalence of microhaemorrhage annotations in smaller regions.

### Differential composition analysis reveals intricate plaque and IPH area composition changes

The additive MIL model uniquely provides continuous IPH measurements, extending beyond binary classification. This allows us, for instance, to investigate the compositional differences between plaques containing model-IPH and those that do not. To this end, we quantified the intensity of all 9 stains in Athero-Express using SlideToolKit^33^. For each stain, we calculated the correlation between the plaque’s stained ratio and the model-IPH area size. In addition, we also directly compared the plaque’s stained ratio in groups between model-IPH-positive and model-IPH-negative regions (**Supplemental Figure 7**). When looking at the correlation between the plaque’s stained ratio and the model-IPH area size, multiple markers vary significantly: We see that SMA (r = -0.30, p-value = 4.53×10^−37^), EVG (r = -0.18, p-value = 1.13×10^−11^), and SR (r = -0.08, p-value = 4.01×10^−3^) have significantly less staining in plaques with an increasing model-IPH area, while GLYCC (r = 0.26, p-value = 2.02×10^−21^) and FIBRIN (r = 0.30, p-value = 1.80×10^−15^) markers positively correlate with model-IPH area (**Figure 5A-I, Supplemental Table 8**).

**Figure 5:**
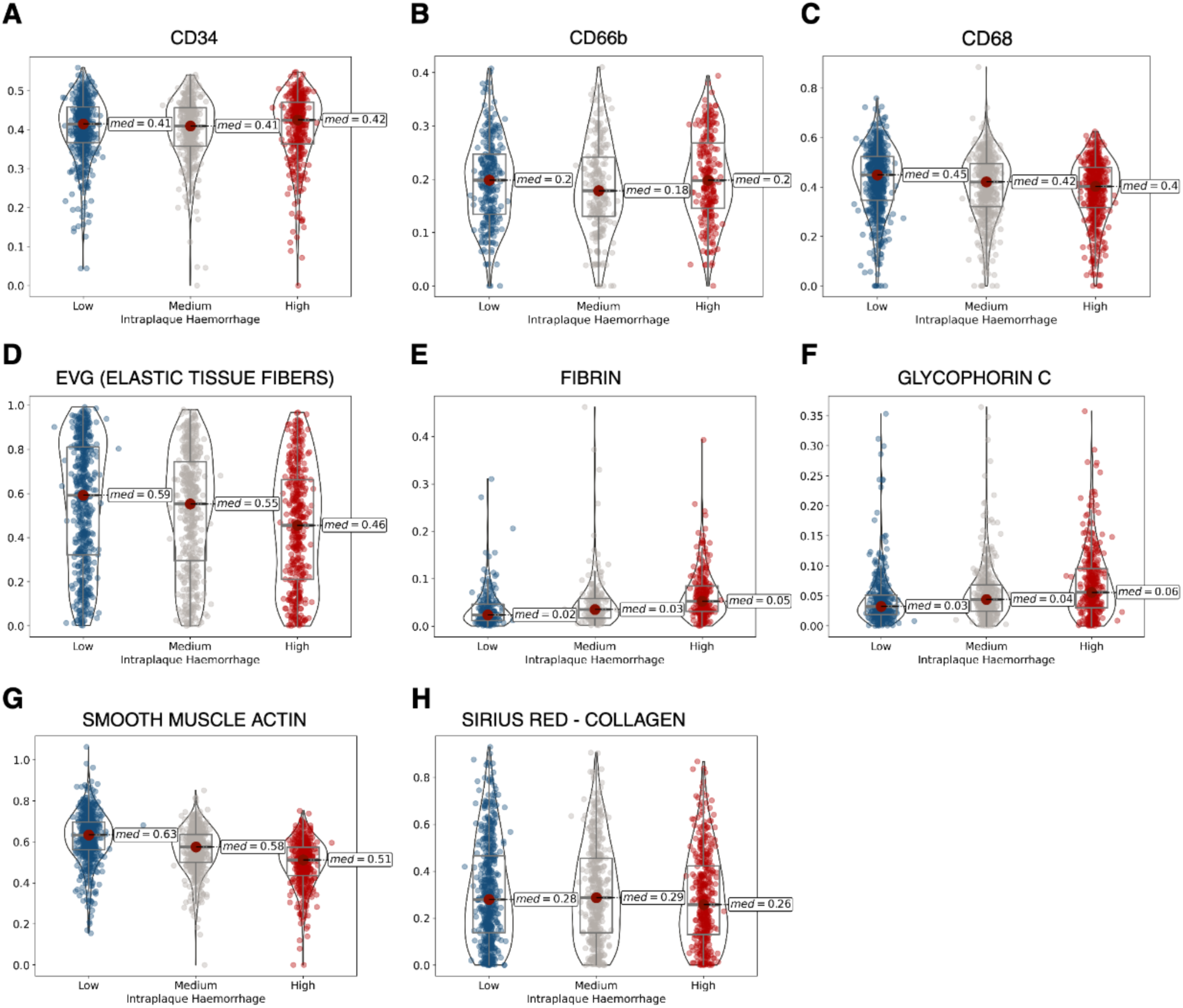
Histological stain quantification across model-IPH area. Violin plots of SlideToolKit^10^ derived biomarkers across different levels of intraplaque haemorrhage (low, medium and high): CD34 (**A**), CD66b (**B**), CD68 (**C**), elastic tissue fibres (**D**), fibrin (**E**), glycophorin C (**F**), smooth muscle actin (**G**) and sirius red (**H**).

When directly comparing model-IPH-positive and model-IPH-negative regions, there are substantial compositional differences, including: less SMA (r = -0.79, p-value = 1.69×10^−234^), H&E (r = -0.49, p-value = 1.09×10^−92^), SR (r = -0.52, p-value = 2.22×10^−87^), CD34 (r = -0.46, p-value = 2.30×10^−63^), EVG (r = -0.33, p-value = 1.87×10^−36^), and CD68 (r = -0.25, p-value = 9.93×10^−25^), whereas there is significantly more GLYCC staining (r = 0.56, p-value = 4.20×10^− 91^) and FIBRIN (r = 0.63, p-value = 5.02×10^−77^). Notably, while model-IPH area positively correlates with manually counted neutrophils at the plaque level (r = 0.41), CD66b staining is reduced within model-IPH-positive regions specifically (r = -0.15, p-value = 6.39×10^−6^), suggesting that these two measures reflect neutrophil biology at different spatial scales (**Supplemental Table 9**).

Collectively, these findings reflect a complex interplay of extracellular matrix (ECM) degradation, increased coagulation and haemorrhage, decrease in smooth muscle cells and neovascularization, and a modest decrease of neutrophil infiltration in areas of IPH.

### Model’s IPH metrics recapitulates known associations with clinical outcome

To determine whether model-IPH metrics recapitulate established clinical associations, we evaluated the potential of our model-IPH metrics to classify preoperative symptom presentation (TIA and stroke). We trained two CatBoost classifiers that included model-IPH metrics, quantified stainings, and clinical metadata. The models classified TIA symptoms with 64% accuracy and stroke with 66%. For both types of symptom presentation, we see that the most predictive features as measured via feature importance scores, are model-IPH area, age, BMI and Glomerular Filtration Rate (GFR) (**Supplemental Figure 5**). Crucially, these models outperformed both the baseline with only age and sex as classification variables (55% accuracy for both TIA and stroke) as well as the baseline with all the clinical metadata (61% and 64% accuracy for TIA and stroke respectively), demonstrating the added value of image-derived phenotypes.

Next, we sought to replicate previously reported findings by Vrijenhoek et. al.^34^, demonstrating a significant, sex-dependent effect of manual-IPH on cardiovascular outcome. Following metadata imputation (mean certainty = 0.903; **Supplemental Figure 6**; **Supplemental Table 10**), we ran a Cox regression using 883 patients (626 males, 257 females). In the overall cohort, both manual-IPH score (HR=1.38, 95% CI: 1.00–1.90, p-value = 0.05) and the model- IPH probability score (HR=1.70, 95% CI: 1.05–2.77, p-value = 0.03) were significantly associated with clinical outcome. When stratified by sex, our model-IPH accurately reproduced known male-specific risk signature^34^. In male patients, our continuous model-IPH probability score highlights greater sensitivity for recovering patients with IPH (HR=1.94, 95% CI: 1.09– 3.46, p-value = 0.02) than both binary model-IPH (HR=1.46, 95% CI: 1.01–2.10, p-value = 0.04) and traditional manual-IPH scoring (HR=1.52, 95% CI: 1.02–2.26, p-value = 0.04). The model-IPH area did not show any significant associations (**Supplemental Figure 8**).

## Genetic and transcriptomic characterisation of IPH variability reveals a macrophage & foam cell signature

To investigate the genetic basis of IPH presence and thereby validating the PHENOMICL model, we conducted a genome-wide association study (GWAS) using both the manual-IPH label and the model-IPH classification (**Supplemental Figure 9)**. Whilst both GWAS resulted in no significant hits, we observed a strong correlation between the beta values of the two models, and a broader range of variants (p-value = 5×10^−6^) for the model-IPH GWAS, indicating that our model recapitulates the same associations as the manual scoring.

To further identify genes associated with model-IPH and its variability, we profiled the transcriptome of n=1,026 plaques and performed differential gene expression (DGE) analyses on both model-IPH abundance and the amount of plaque biomarkers derived from 8 concurrent Immunohistochemistry (IHC) stains. The DGE analysis of the model-IPH area identified 1,412 significant (FDR 5%, p-value < 0.05) differentially expressed genes (854 highly expressed, 558 low expressed).

Furthermore, we looked for associations between differentially expressed genes and atherosclerosis-related outcomes in the literature. *TIMP3* and *MMP9* are associated with increased model-IPH area. It is known that the balance between *TIMP3* (an inhibitor of matrix metalloproteinases) and MMPs plays a critical role in the stability of the ECM, whose disruption can lead to adverse cardiovascular events^35–37^. In Open Targets^38^, *MMP9* is reported to be associated with increased triglycerides and decreased HDL cholesterol levels^39^. Several of the identified differentially expressed genes (*ATP2B1*, *CACNB2*, *LPA*, *LSP1* and *TRIB1*) have also been previously prioritised as the likely targets in a GWAS for cardiovascular disease^40^. (**Supplemental Table 11**).

Functional enrichment revealed pronounced activation of transcriptomic factor (TF) networks associated with high model-IPH areas. Among these were *JUN* (p-value = 1.05×10^−37^), *SP1* (p-value = 2.65×10^−32^), *NFkB* (p-value = 1.05×10^−30^). Similar results were obtained for GLYCC. Conversely, these TFs were mostly inactive in CD34, CD68 and SMA (**Figure 6D**). *JUN*, *SP1* and *NFkB* all play critical roles in regulating genes involved in inflammation (cytokine response, leukocyte recruitment, foam cell formation and activation), vascular cell adhesion, ECM remodelling and oxidative stress, processes that are all critical for sustaining a local inflammatory response and contribute to the destabilisation of the plaque microenvironment.

**Figure 6:**
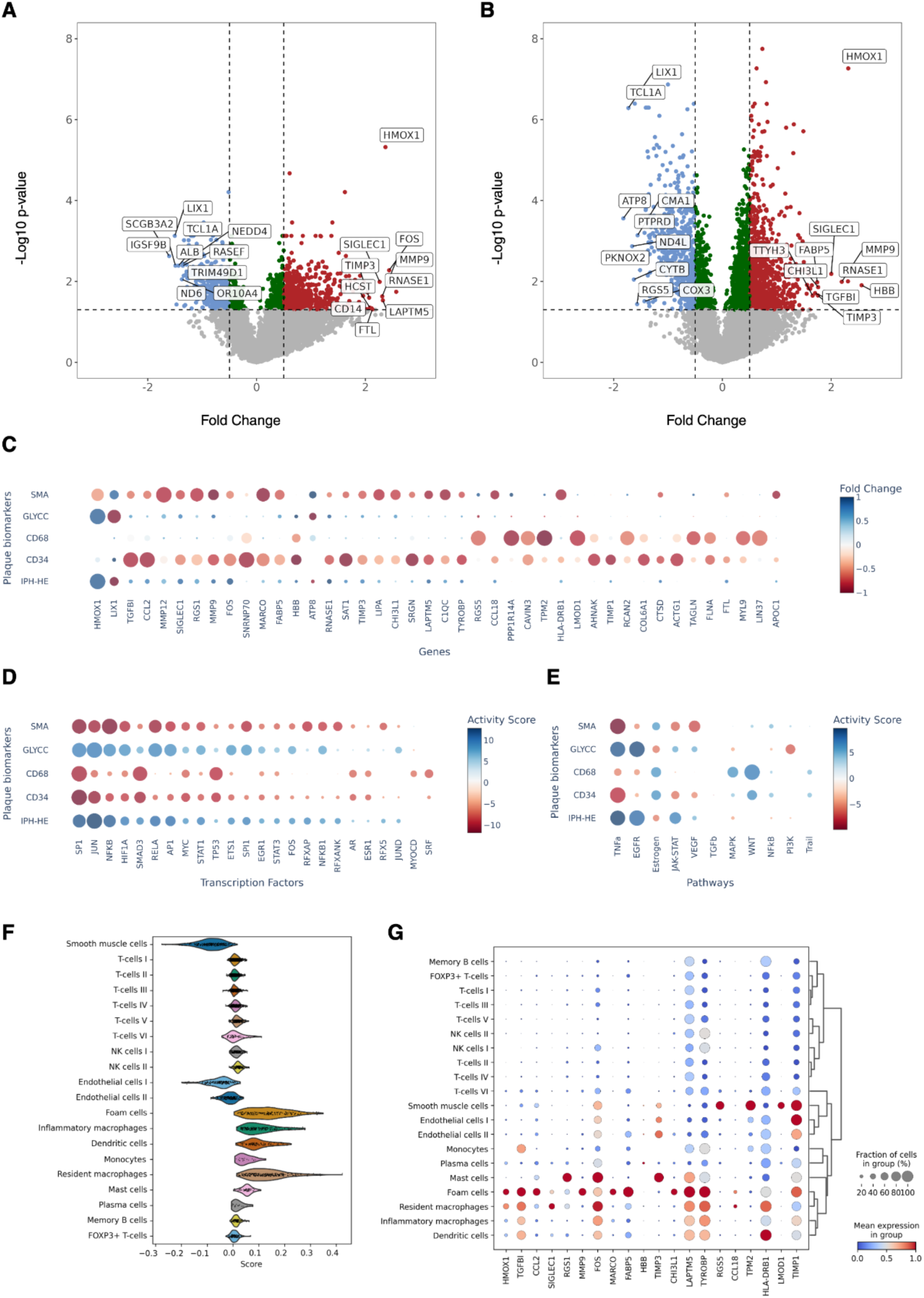
Differential gene expression and downstream analysis. Volcano plots of the differential gene expression using the model-IPH area (**A**) and the amount of glycophorin C (**B**)as continuous phenotypes. (**C**) Top 10 differentially expressed genes per phenotype, ordered by total absolute fold change; the color indicates the fold change, while the dot size correlates with its significance. (**D**) Transcription factor and (**E**) pathway activity score per phenotype; in both cases, the colour of the dot represents the activity score, whereas its size indicates the significance of the association. (**F**) Violin plot of the set of model-IPH area-associated genes scored across the cell types of a single-cell reference; the plot clearly shows a positive enrichment of foam cells, inflammatory and resident macrophages, and a negative enrichment of smooth muscle cells. (**G**) Mean expression of the top 20 genes reported in panel (C)per cell type; the colour of the dot depicts the mean expression value and its size denotes the fraction of cells expressing the gene.

These enrichments were further corroborated by pathway activity inference analyses, demonstrating a large positive enrichment for TNF-α (p-value = 1.83×10^−12^) and EGFR (p-value = 1.83×10^−12^), whilst a negative enrichment for estrogen (p-value = 1.0×10^−6^) and PI3K signaling (p-value = 1.80×10^−5^) (**Figure 6E**). TNF-α is a critical master regulator of inflammation and leukocyte recruitment into the plaque^41^, whilst estrogen has been shown to have protective vascular effects in women^42^. Functional enrichment of biological terms with MSigDB revealed that many of our differentially expressed (DE) genes have a significant overlap with several gene sets, like heart macrophages markers (FDR p-value = 1.70×10^−38^), innate immune response (FDR p-value = 2.46×10^−14^), regulation of leukocyte adhesion to arterial endothelial cells (FDR p-value = 2.03×10^−2^) and eosinophil activation and chemokine binding (FDR p-value = 6.28×10^−5^).

When we mapped the model-IPH genes onto a single-cell RNA-sequencing dataset, we confirmed that these differentially expressed genes are enriched in foam cells, inflammatory and resident macrophages, while being depleted in smooth muscle cells (**Figure 6F-G**), confirming our previously observed negative correlation between model-IPH and SMA and increased performance in detecting model-IPH when adding CD68 and EVG stains to the model.

In summary, we derived an expression signature of model-IPH and other image-derived plaque biomarkers through differential expression, transcription factor and pathway activity analyses and gene scoring in a single cell reference, demonstrating the importance of ECM stability and the important role of foam cells. This DGE analysis shows that our model-IPH area captures biologically meaningful regions of the plaque, characterized by inflammatory activation, ECM remodelling, and macrophage-driven processes consistent with known IPH pathology.

### Spatial transcriptomics localizes IPH-associated gene signatures to macrophage-enriched haemorrhagic regions

To validate the bulk-transcriptomic findings and link model-IPH-associated gene expression to specific tissue regions and cell types in a spatial context, we performed spatial transcriptomics profiling using Visium HD (10x Genomics) on 8 carotid plaque samples. We spatially mapped the top 10 upregulated model-IPH genes, which demonstrated robust, spatial concordance with PHENOMICL heatmaps and GLYCC staining (**Figure 7A**). In addition, we assigned the most prominent cell type to each spatial bin using cell type specific gene signatures derived from our single-cell RNA-sequencing data, enabling spatial deconvolution. Consistent with our single-cell analysis, regions with high model-IPH gene expression were further enriched for macrophage-like cell type signatures. In contrast, areas with smooth muscle cell-like signatures show negligible model-IPH gene expression or overlap with PHENOMICL heatmaps.

**Figure 7:**
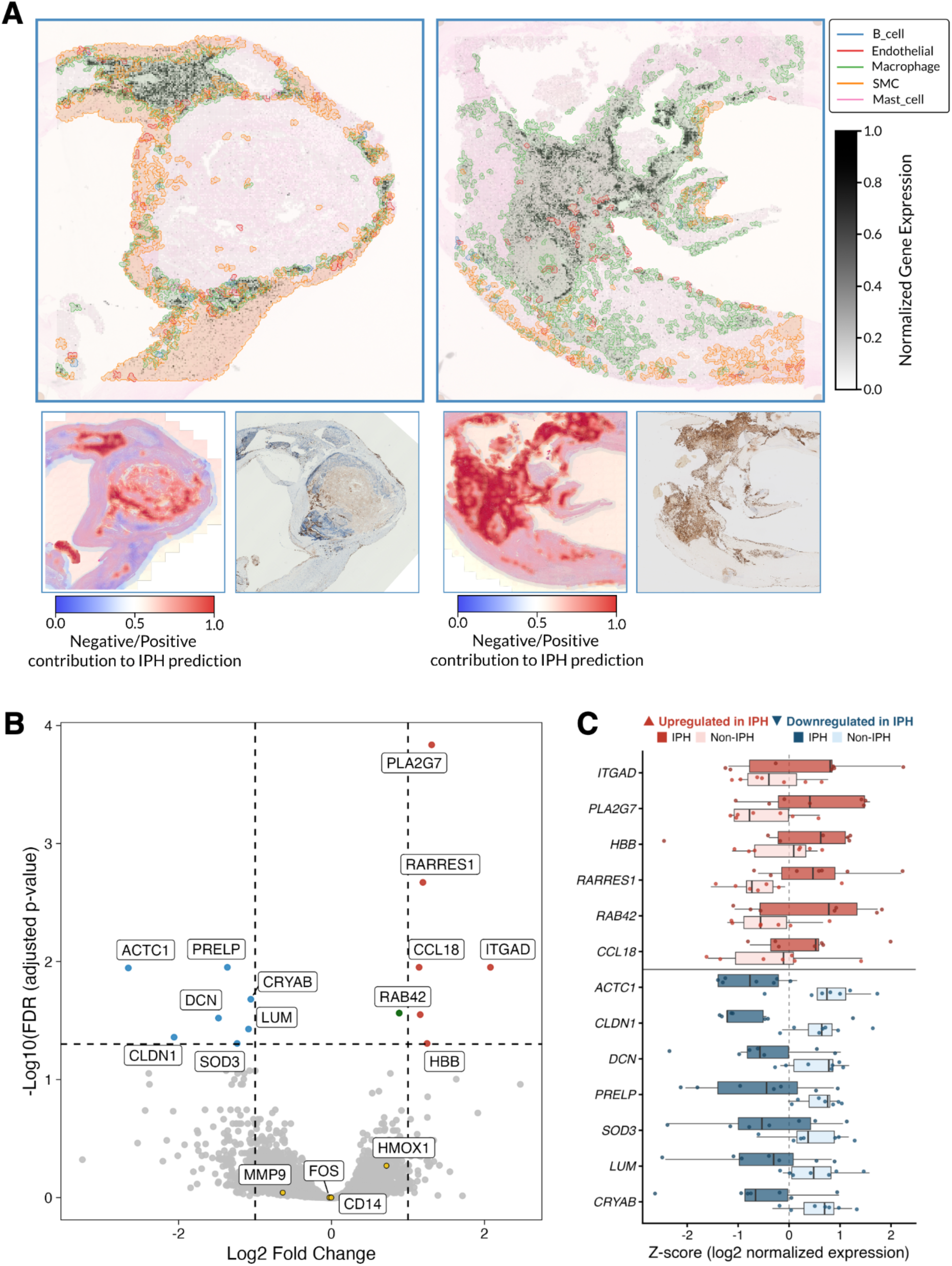
Spatial transcriptomics validates model-IPH upregulated signatures. Top 10 model-IPH upregulated genes show expression in macrophage-like cell regions. Additional model-IPH heatmaps and GLYCC stains are shown (**A**). Volcano plot of the spatial transcriptomic differential gene expression between regions of IPH and non-IPH (**B**). Gene expression between IPH and non-IPH regions of the 13 differentially expressed genes (**C**).

Spatial pseudobulk DGE conducted between model-IPH positive and negative regions identified 13 significantly differentially expressed genes (6 upregulated, 7 downregulated; FDR 5%, p-value < 0.05; **Figure 7B-C**). Regions annotated as model-IPH positive showed increased expression of hemoglobin-associated genes (e.g. *HBB*), reflecting erythrocyte infiltration and hemoglobin deposition^43^ alongside enrichment of inflammatory and lipid-signaling leukocyte markers, including *PLA2G7*, *ITGAD*, and *CCL18*^44–47^.

Simultaneously, model-IPH positive regions show reduced expression of structural genes. Extracellular matrix regulators (*DCN*, *LUM,* and *PRELP*)^48,49^ were downregulated, reflecting broader alterations in matrix organization associated with unstable plaques. Additionally, reduced expression of endothelial adhesion markers (e.g. *CLDN1*)^50^ supports the presence of a compromised and leaky microenvironment. Furthermore, decreased expression of oxidative stress response genes (*SOD3*, *CRYAB*) suggests a reduced capacity to counteract oxidative stress within the haemorrhage regions^51,52^. Consistent with bulk transcriptomic and IHC findings, *HMOX1* showed a positive fold change in model-IPH positive regions (log2FC = 0.72, nominal-p = 0.021), though this did not reach FDR-corrected significance (padj = 0.54), likely reflecting limited statistical power in the spatial dataset (n=7).

Together, these findings indicate that PHENOMICL not only captures morphological features, but identifies regions in the plaque that reflect the expected molecular and cellular microenvironment of IPH. The spatial gene expression patterns capture established features of IPH, including inflammation, oxidative stress, and extracellular matrix destabilization, supporting the validity of the models IPH localization and biological relevance. This provides direct in-tissue validation of the spatial accuracy of the model and supports its ability to localize biologically meaningful plaque features.

### Cell-cell communication analyses implicate ACKR1-mediated endothelial-macrophage interactions as drivers of intraplaque haemorrhage

To identify the cellular networks driving microvascular instability in IPH, we performed cell-cell communication analysis using MetaPlaq v2^53^ an integrative atlas of atherosclerosis (**Figure 8**). Comparing early and advanced lesions revealed differential regulation of various ligand-receptor interactions, specifically involving the endothelial receptor ACKR1, a chemokine receptor we found to be significantly upregulated in the model-IPH DGE. Endothelial ACKR1 expression is implicated in macrophage polarization during aortic dissection and is induced upon the exposure to whole blood^54,55^. Accordingly, advanced plaques showed upregulation of macrophage chemokines ligands to ACKR1^high^ endothelial cells. The TREM2^high^ /PLIN2^high^ lipid associated macrophage (LAM) CCL2 and CXCL8 (ligand) to ACKR1^high^ endothelial cell ACKR1 (receptor) interaction was significantly upregulated (**Figure9B-D**). CCL2 acts as a master regulator of macrophage recruitment^56^ and promotes neovascularization through PI3K/AKT signalling in models of diabetic wound healing and gastric cancer^57,58^. Aberrant neovascularization, which increases in plaque progression, has been proposed as a causal mediator of intraplaque haemorrhage due to incompetency of newly formed microvessels^59–61^. Notably, we identified upregulation of tissue-resident macrophage CCL18 (ligand) with activated endothelial cell ACKR1 (receptor). CCL18 colocalizes with CD68 and CD163 (a marker of heme-loaded macrophages^62^) in ruptured plaques. However, its direct interaction with ACKR1 has not been investigated in IPH and atherosclerosis. This implicates the CCL- ACKR1 axis as a potentially novel mediator of angiogenesis and associated intraplaque haemorrhage.

**Figure 8:**
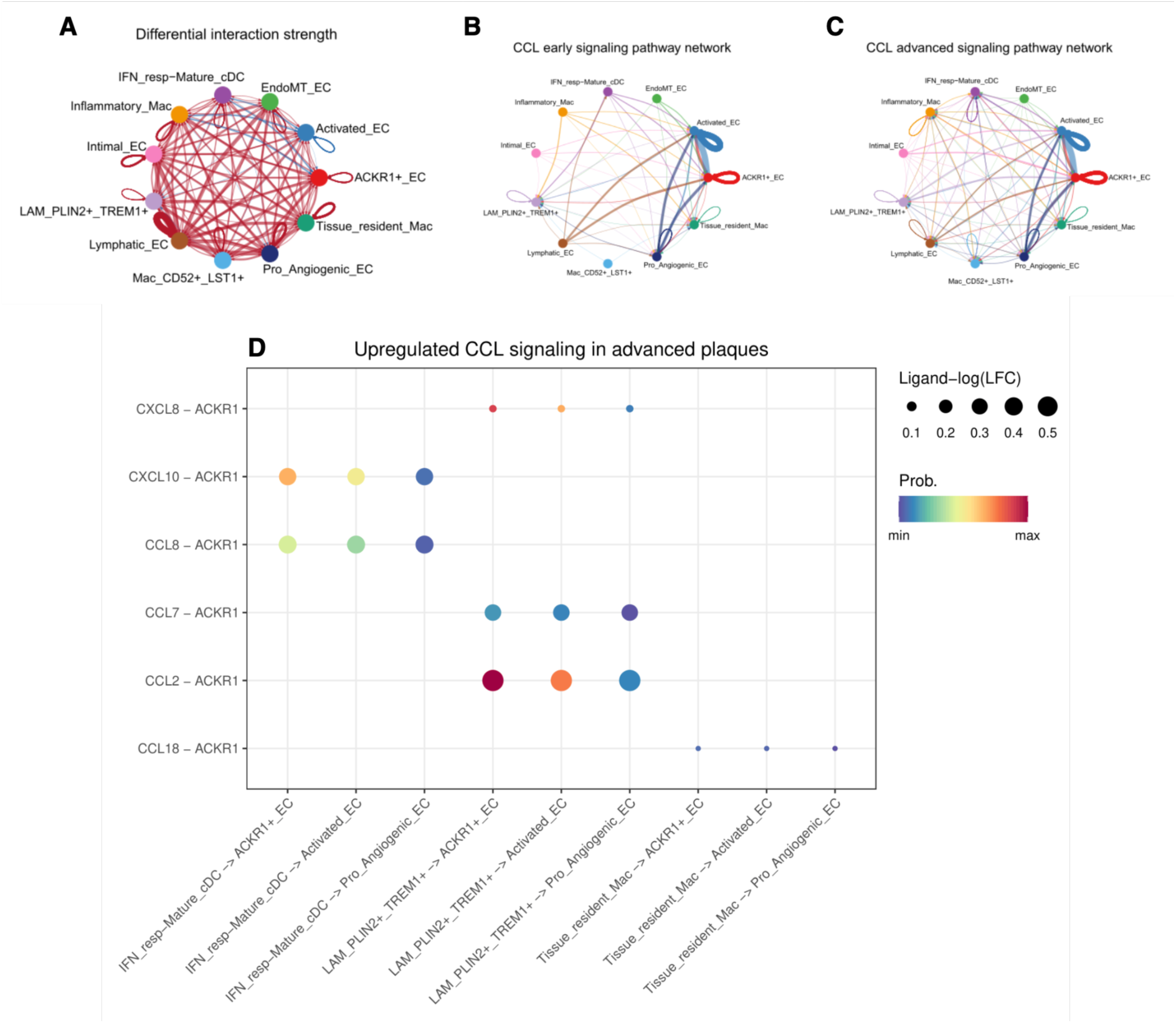
CCL-ACKR1 axis is potential driver of intraplaque haemorrhage. **(A)** Circle plot showing overall differential interaction strength in advanced vs early lesions for macrophage and endothelial cell subtypes. **(B & C)** CCL signaling network activity in early **(B)** and advanced (**C)** atherosclerotic lesions. **(D).** CCL-ACKR1 differentially upregulated in advanced atherosclerotic lesions.

## Discussion

This study represents the largest digital pathology analysis of intraplaque haemorrhage (IPH) to date, leveraging 2,595 carotid plaques from the Athero-Express Biobank. We developed PHENOMICL, an attention-based additive multiple instance learning (MIL) framework, enabling scalable histological quantification and spatial localization of IPH. This spatial detection enables downstream integration with molecular and clinical data at scale, a workflow that remained impractical using traditional manual methods.

Automated quantification of IPH using the standard H&E histology alone achieved the highest classification performance (AUROC = 0.86), suggesting that PHENOMICL can be effectively applied using the most widely available staining protocol. The incorporation of CD68 or EVG showed significant performance gains, reaching an AUROC of 0.92. Robust validation across femoral arteries and independent cohorts (HeCES2 and AtherOMICS) demonstrates PHENOMICL’s generalizability. In the fibrotic, calcified femoral vascular bed, structural stains (EVG and SR) slightly outperformed H&E, highlighting the importance of extracellular matrix (ECM) integrity in plaque characterization. Conversely, cell-specific stains, like GLYCC and SMA, likely highlight their high specificity which limits its ability to capture broader haemorrhage-related plaque composition features. External validation in the HeCES2 cohort, an analogous carotid plaque cohort, confirmed the robustness of PHENOMICL. Unlike the Athero-Express, where manual-IPH is scored solely from the histological WSI, the HeCES2 cohort uses a composite score incorporating macroscopic morphology. Meaning, macroscopic IPH observed outside the histological WSI still contributes to a positive label. This introduces a slide-label mismatch most prominent in cases with small IPH. Similarly, spatial localization validation using the AtherOMICS cohort showed strong cohort-level correlation between model-derived and manually annotated IPH area. However, patch-level spatial agreement revealed that smaller IPH regions are harder to identify. Specifically, small microhaemorrhages are vulnerable to boundary-assignment disagreements, where IPH near patch borders is differently assigned by the model versus the manual annotation. Additionally, manual annotations often include “empty spaces” within the broader IPH region, which PHENOMICL, lacking global context, excludes as non-tissue.

Consistent with previous studies^34,63^, model-IPH emerged as a key phenotype linked to plaque instability and adverse clinical outcomes, including stroke and major adverse cardiovascular events (MACE). Our model-IPH metrics showed stronger predictive power compared to manual-IPH labelling, particularly in male patients. Beyond IPH, quantified histological stainings collectively reflect a complex interplay of extracellular matrix (ECM) degradation, increased coagulation and haemorrhage, decrease in smooth muscle cells and neovascularization, and a modest decrease of neutrophil infiltration in areas of IPH. The divergent neutrophil findings across spatial scales - a positive correlation between model-IPH area and neutrophil count alongside reduced CD66b staining within haemorrhagic regions - likely reflect two distinct biological phenomena. At plaque level neutrophil recruitment may be driven by the systemic inflammatory response associated with greater IPH burden, while the regional decrease of CD66b in haemorrhage areas could reflect processes such as NETosis, apoptosis, or efferocytosis by macrophages. Furthermore, incorporating model-IPH metrics and quantified stainings with clinical data achieved greater symptom classification compared to baseline models, demonstrating the utility of image-derived phenotypes in understanding the role of plaque composition in clinical presentations.

Our bulk and spatial transcriptomic analyses confirmed that PHENOMICL identifies regions displaying transcriptional signatures associated with IPH, shedding light on the biological mechanisms underpinning plaque vulnerability. The bulk transcriptomics showed an upregulation of key genes such as *TIMP3* and *MMP9*, reflecting the critical balance between extracellular matrix stability and degradation^35–37^. This, in combination with spatial downregulation of structural matrix genes (*DCN*, *LUM*, and *PRELP*), provides a clear molecular signature of active extracellular matrix degradation. Furthermore, bulk- (*CXCL1*, *CXCL4*, and *SIGLEC1*) and spatial (*PLA2G7*, *ITGAD*, and *CCL18*) genes linked to inflammation highlight additional inflammatory pathways driving plaque instability. Enrichment analyses identified TNF-α activation and estrogen suppression as central pathways, aligning with the structural and inflammatory features of plaques^41,42^. These molecular insights corroborate the strong classification performance of structural stains like SR and EVG and further support the validity of the model’s localization capabilities, linking histological findings to transcriptional activity.

Cell-cell communication analysis revealed deeper insight into the mechanistic drivers behind IPH. We observed several interactions involving endothelial ACKR1 (receptor) and macrophage chemokines ligands, with the strongest signal arising from TREM2^high^ /PLIN2^high^ lipid associated macrophage CCL2 signaling to ACKR1^high^ endothelial cell. CCL2 is a master regulator of macrophage recruitment and secretion has been shown to promote neovascularization. A potential novel interaction was also observed between tissue resident macrophage CCL18 (ligand) with activated endothelial cell ACKR1 (receptor), which has not been investigated in relation to IPH and atherosclerosis. CCL18 has been shown to colocalize with the macrophage marker CD68 and CD163, a marker of heme-loaded macrophages, hinting at a possible link to haemorrhage associated macrophage activity.

Despite the advancements, several limitations must be acknowledged. While we identified sex-based differences in symptom classification, with model-IPH showing stronger predictive associations in males, this study did not stratify analyses by sex comprehensively. Additionally, the absence of image registration between serial stained plaque sections limited the ability to fully integrate stain-specific information. Future studies should address these limitations by incorporating sex-stratified analyses and refining data integration methods. Nevertheless, PHENOMICL sets the stage for future integration of image-derived phenotypes with clinical data and (spatial) transcriptomics. Due to its generalizability, PHENOMICL can be extended to other phenotypes and diseases, providing a robust, interpretable framework for understanding the pathophysiology of atherosclerosis and deepening our understanding of plaque biology.

## Methods

### Athero Express Biobank

#### Study design

Carotid and femoral atherosclerotic plaques were obtained from patients undergoing arterial endarterectomy and included in the Athero-Express Biobank Study (AE), an ongoing biobank study at the University Medical Centre Utrecht (Utrecht, The Netherlands) and the St. Antonius Hospital (Nieuwegein, The Netherlands)^29^. The medical ethical committees of the respective hospitals approved the AE which was registered under number 22/088. This study complies with the Declaration of Helsinki, and all participants provided written informed consent.

#### Sample collection

Blood samples were obtained prior to surgery and stored at -80℃. Carotid plaque specimens were removed during surgery and immediately processed in the laboratory following standardized protocols. Specimens were cut transversely into segments of 5 mm^29^. The culprit lesion (the region with the most severe stenosis) was identified, fixed in 4% formaldehyde, embedded in paraffin, and processed for histological examination^29^. The remaining segments were stored at -80℃.

#### Clinical characteristics

Various clinical characteristics are described throughout the paper, with additional details provided in **Supplemental Table 12.** The composite endpoint mentioned includes any death of presumed vascular origin (fatal stroke, fatal myocardial infarction, sudden death, other vascular death), nonfatal stroke, nonfatal myocardial infarction, and any arterial vascular intervention that had not already been planned at the time of inclusion (eg, carotid surgery or angioplasty/stenting, coronary bypass, percutaneous coronary intervention, peripheral vascular surgery or angioplasty/stenting).

#### Histological staining

We described the standardized (immuno)histochemical analysis protocols used in the AE before^3,29^. In short, 10-micron cross-sections of the paraffin-embedded segments were cut using a microtome, stained using standardized protocols, and examined under a bright-light microscope. The following stainings are performed to characterize the plaque: Picro Sirius red (SR, collagen and fat using polarized light), CD68 (macrophages), smooth muscle cell alpha-actin (SMA, smooth muscle cells), hematoxylin and eosin (H&E, thrombus and calcifications and elastin), Verhoeff’s Van Gieson (EVG, elastic fibres), FIBRIN (fibrin deposits and blood clots), glycophorin C (GLYCC, red blood cells), CD34 (endothelial cells), and CD66b (neutrophils). The immunohistochemical staining for CD68 and SMA stains are performed fully automated (Ventana Medical Systems, Tucson, Arizona) (**Supplemental Table 13**).

Intraplaque haemorrhage (IPH) was scored as no/yes using a hematoxylin and eosin staining (H&E) as described before^10^. All histological observations were performed by the same dedicated technician and interobserver analyses have been reported previously^64^.

#### Whole-slide scanning

We set up ExpressScan to obtain whole-slide images (WSIs) by scanning stained slides at 40x using a Roche Ventana iScan HT or Hamamatsu C12000-22 Digital slide scanner. WSIs were stored digitally as z-stacked .TIF (Roche) at 0.25 micron/pixel or .ndpi (Hamamatsu) at 0.23 micron/pixel brightfield microscopy images^10^.

#### Bulk RNA isolation and sequencing

A total of 1,093 plaque segments were selected from patients included in the Athero-Express Biobank Study between 2002 and 2017 in two subsequent experiments, Athero-Express RNA Study 1 and 2 (AERNAS1: n=622, 2002–2016; AERNAS2: n=471, 2002–2017)^65^. The atherosclerotic plaques were grounded while frozen with liquid nitrogen, and, after that, TriPure (Roche, Cat# 11667165001) was added, and the plaque pieces were further disrupted by a Precellys 25 homogenizer (Bertin Instruments). The sample was incubated at room temperature for 5 minutes and centrifuged at 20,000g for 1 minute at 4℃. The supernatant was mixed with chloroform and incubated at room temperature for 15 minutes. The sample was centrifuged at 12,000×g for 5 minutes at 4°C, and the upper phase was used for RNA isolation. Then, isopropanol and GlycoBlue (Invitrogen, Cat# 10301575) were added to the aqueous phase to precipitate the RNA and centrifuged at 12,000g for 10 minutes at 4°C. The pellet was washed with 75% ethanol and resuspended in RNase-free water.

The RNA isolated from the archived advanced atherosclerotic lesion is fragmented. They have, therefore, employed the CEL-seq2 library preparation protocol^66^. CEL-seq2 yielded the highest mappability reads to the annotated genes compared to other library preparation protocols. The methodology captures the 3′ end of polyadenylated RNA species and includes unique molecular identifiers (UMIs), which allow direct counting of unique RNA molecules in each sample. Then, 50 ng of total RNA was precipitated using isopropanol and washed with 75% ethanol. After removing ethanol and air-drying the pellet, a primer mix containing 5 ng of primer per reaction was added, initiating primer annealing at 65°C for 5 minutes. Subsequent reverse transcription reaction was performed: first-strand reaction for 1 hour at 42°C; heat-inactivated for 10 minutes at 70°C; second-strand reaction for 2 hours at 16°C; and then put on ice until proceeding to sample pooling. The primer used for this initial reverse transcription reaction was designed as follows: an anchored polyT, a unique 6-bp barcode, a UMI of 6 bp, the 5′ Illumina adapter and a T7 promoter, as described. Each sample now contained its own unique barcode due to the primer used in the RNA amplification, making it possible to pool together complementary DNA (cDNA) samples at seven samples per pool. cDNA was cleaned using AMPure XP beads (Beckman Coulter, Cat# A63882), washed with 80% ethanol and resuspended in water before proceeding to the in vitro transcription (IVT) reaction (Thermo Fisher Scientific, Cat# AM1334) and incubated at 37°C for 13 hours. Next, primers were removed by treating with Exo-SAP (Affymetrix, Thermo Fisher Scientific, Cat# 78201.1.ML), and amplified RNA (aRNA) was fragmented and then cleaned with RNAClean XP (Beckman-Coulter, Cat# A63987), washed with 70% ethanol, air-dried and resuspended in water. After removing the beads using a magnetic stand, RNA yield and quality in the suspension were checked by Bioanalyzer (Agilent Technologies).

cDNA library construction was then initiated by performing a reverse transcription reaction using SuperScript II reverse transcriptase (Invitrogen/Thermo Fisher Scientific, Cat# 18064022) according to the manufacturer’s protocol, adding randomhexRT primer as a random primer. Next, polymerase chain reaction (PCR) amplification was done with Phusion High-Fidelity PCR Master Mix with HF buffer (New England Biolabs, F531L) and a unique indexed RNA PCR primer (Illumina) per reaction, for a total of 11–15 cycles, depending on aRNA concentration, with 30 seconds of elongation time. PCR products were cleaned twice with AMPure XP beads (Beckman Coulter, Cat# A63882). Library cDNA yield and quality were checked by Qubit fluorometric quantification (Thermo Fisher Scientific, Cat# Q32851) and Bioanalyzer (Agilent Technologies), respectively. Libraries were sequenced on the Illumina NextSeq 500 platform, paired-end, 2 x 75 bp.

#### Single-cell RNA isolation and sequencing

In total 46 atherosclerotic plaque segments were used for scRNA sequencing and isolation described in detail by Depuydt *et al.* ^67^ and Slenders *et al.*^68^. In summary, all samples were collected from patients undergoing a carotid endarterectomy procedure. To examine the transcriptome, the plaque samples were enzymatically dissociated, filtered, and cryopreserved for subsequent fluorescence-activated cell sorting (FACS) and single-cell RNA sequencing (scRNA-seq). Viable cells were identified using Calcein AM and Hoechst staining before being sorted on a Beckman Coulter MoFlo Astrios EQ. Individual cells were dispensed into 384-well plates, lysed, and processed following the SORT-seq protocol, incorporating CEL-Seq2^66^ library preparation and sequencing on an Illumina NextSeq 500. scRNA-seq data processing involved stringent quality control, normalization using Seurat^69^, and clustering via canonical correlation analysis (CCA) with subsequent tSNE visualization. Cell type annotation was performed using differential gene expression analysis against BLUEPRINT reference datasets^70^, and pathway analysis was conducted using EGSEA^71^ with hallmark gene sets from GSEA. Clustering of cell populations provided a high resolution of the cellular landscape in atherosclerotic lesions, further distinguishing macrophages, lymphocytes, mast cells, and endothelial cells.

#### MetaPlaq

For the cell type labeling of the spatial transcriptomics data, we used data from MetaPlaq, a multimodal atlas of human atherosclerotic arterial beds comprising over a million cells across single-cell transcriptomics from atherosclerotic plaques^53^. Single cell RNA-seq data from coronary, carotid, femoral and tibial arteries was collected and processed as detailed in Mosquera *et al*. (2026)^53^. Briefly, after aligning raw sequencing counts to the hg38 reference genome, each library counts matrix was preprocessed using a QC pipeline implemented within the scRNAutils R package (https://github.com/MillerLab-CPHG/scRNAutils/tree/dev). Within this pipeline, we first identified doublets by performing 3 iterations of artificial doublet generation and detection with scDblFinder^72^ (v.1.16.0) and consensus doublets across all iterations were removed from downstream analyses. To correct for ambient RNA, we ran DecontX^73^ within the celda R package (v.1.18.2) in doublets-filtered raw counts matrices using default parameters.

To account for the extensive heterogeneity in terms of overall data quality and sequencing depth, we further filtered cells using an adaptive thresholding approach. First, we build a full rank matrix of cells by QC covariates (% Mitochondria, %Ribosomal reads, N Reads and N Genes). We then compute an outlyingness score using the robustbase R package (v0.99.7) with the **adjOutlyingness** function. Each cell receives an outlyingness score and then we remove outliers based on the Median Absolute Deviation (MAD) using the **isOutlier** function implemented within the scater R package^74^ (v1.30.1). We chose a rather stringent MAD threshold since higher MADs led to an increased proportion of low-quality cells. Because this feature is quite sparse, we also remove cells with > 1% reads mapped to hemoglobin genes as these are likely contaminating erythrocytes.

### Post-integration Quality Control and Level 1 annotations

Based on the benchmark detailed in Mosquera *et al.* (2026)^53^, we used the scArches reference mapping framework^75^ to harmonize libraries across multiple arterial beds and sequencing platforms (10X, Cel-seq2 and Smart-seq2). We used the scArches latent space for computing a kNN graph with the function **scanpy.pp.neighbors**, which was used as input for cell clustering with the Leiden algorithm using a resolution of 1.5. We then performed a first pass of manual annotations based on canonical markers and refined labels by cross-referencing our annotations against scArches predicted labels from the Mosquera *et al.* (2023)^76^ reference. We flagged problematic clusters that showed a mixture of predicted labels, were dispersed across multiple lineages and/or contained abnormally low prediction confidence scores as these likely depicted remaining doublets. Upon removing low-quality clusters, we did a second pass of annotations to establish the final L1 labels.

#### DNA isolation and genotyping

The Athero-Express genotyping has previously been described in detail^12,77^. In short, DNA was extracted from 2,652 EDTA blood or (when no blood was available) plaque samples of 2,266 consecutive patients and genotyped in 3 batches irrespective of artery type. Note that 1 patient can have multiple samples, thus we work with a unique patient identifier (UPID) which should not be confused with nor is it linked to a hospital ID, and a sample identifier (studynumber). In the first Athero-Express Genomic Study (AEGS1), 822 patients between 2002 and 2007 were included and genotyped (440,763 markers) using the Affymetrix Genome-Wide Human SNP Array 5.0 (SNP5) chip (Affymetrix Inc., Santa Clara, CA, USA). In the second Athero-Express Genomic Study (AEGS2), 1,022 patients between 2002 and 2013 were included (not overlapping with AEGS1) and genotyped (587,351 markers) using the Affymetrix Axiom® GW CEU 1 Array (AxM). For the Athero-Express Genomics Study 3 (AEGS3), 658 patients between 2002 and 2016, were included and genotyped (693,931 markers) using the Illumina GSA MD version 1 BeadArray (GSA) at the Human Genomics Facility (HUGE-F in Rotterdam, The Netherlands).

After genotyping, we followed community standard quality control and assurance (QC) procedures to clean the whole-genome data obtained in AEGS1, AEGS2, and AEGS3^78^. In the first step, samples that failed genotyping (due to hybridization) and with low average genotype calling (based on manufacturer recommendations) were excluded. Samples with other issues (patient withdrew from the study), sex discrepancies based on available clinical data, relatedness (pi-hat > 0.20) where none should exist, and inbreeding and contamination (average heterozygosity rate ± 3.0 s.d.) were excluded. Samples that turned out to be within (same study number and UPID) or between (different study number but same UPID) related (multiple) pairs were retained. After these initial sample QC steps, variants were filtered out based on positional issues compared to the reference (1000G phase 3, version 5, b37)^79^, duplicate variants IDs, duplicate variant positions, multi-allelic variants, and Hardy-Weinberg Equilibrium (HWE p-value < 1.0×10^−3^). After sample and SNP QC, 780 samples (535 males, 245 females) and 409,023 SNPs in AEGS1, 900 samples (605 males, 295 females) and 529,193 SNPs in AEGS2, and 587 samples (407 males, 180 females) and 661,053 SNPs in AEGS3 remained and were included for imputation.

These 2,267 samples (1,547 males, 720 females) and 1,416,258 unique variants across the three datasets were further pre-processed and taken forward for imputation against TOPMed b38 (R3) using TOPMed Imputation Server (https://imputation.biodatacatalyst.nhlbi.nih.gov/) which includes 445,600,184 variants across chromosome 1–22 and X from 133,597 people^80^.

After imputation we performed principal component analysis to assess the population stratification against 1000G phase 3 data (version 5, including, b38)^79^; 88 samples project to non-EUR populations, and 20 samples project to non-NL populations with EUR. Finally, we randomly picked 2nd and 1st-degree relatives, replicates, or duplicates and post-imputation QC 2,054 samples (1,398 males, 656 females) remain (1,989 NL/EUR, 59 non-EUR, 6 non-NL) covering 73,603,379 unique variants across chromosome 1–22 and X, 1,436 Y-variants and 193 MT-variants (**Supplemental Figure 11**).

### Helsinki Carotid Endarterectomy Study 2 (HeCES2)

To evaluate MIL model performance in a second external dataset, we used H&E sections of carotid plaques from the Helsinki Carotid Endarterectomy Study 2 (HeCES2). HeCES2 is an observational prospective cohort study conducted at Helsinki University Hospital (HUH), Finland, in collaboration with the departments of neurology, vascular surgery, radiology, and pathology. The study protocol has been published elsewhere^81^. The study was approved by the Ethics Committee of medicine of the Hospital District of Helsinki and Uusimaa, and all study patients gave written informed consent. An overview of the cohort characteristics can be found in **Supplemental Table 14.**

Plaque samples from consenting patients with carotid atherosclerosis were collected during carotid endarterectomy. After surgical removal, specimens were rinsed gently with saline, weighed, and photographed. Each specimen was divided into longitudinal slices, of which one representative slice near the most vulnerable area was reserved for histological analysis. This slice was fixed in 10% formalin for 2–4 days, decalcified in EDTA solution for 1–4 weeks depending on the degree of calcification, dehydrated, and embedded in paraffin. 4mm thick sections from paraffin-embedded specimens were stained with Hematoxylin-Eosin (H&E) using an automated staining machine and digitized using a Pannoramic 250 slide scanner (3DHISTECH, Budapest, Hungary) at 20× magnification (0.24 µm/pixel) as whole-slide images in MRXS format.

Histological scoring was performed on one representative longitudinal slice per specimen. All samples were independently graded by two investigators, including a vascular pathologist, following a consensus grading protocol established across 30 pilot specimens. Characteristics linked to plaque instability, including IPH, ulcerations, thrombi, fibrous cap rupture, and luminal irregularity, were assessed in detail. Macroscopic observations were integrated into the histological scoring: features identified macroscopically but absent from the histological section were nonetheless included in the final grade. IPH was quantified as a percentage of total plaque area, with red blood cell accumulations disrupting the plaque architecture identified as haemorrhage. Plaques were classified as IPH-positive if intraplaque haemorrhage was present on either macroscopic or histological assessment. For this study, 485 H&E whole-slide images with corresponding IPH labels were provided. 7 H&E slides were excluded due to missing IPH labels, leaving 478 H&E slides for external validation.

### AtherOMICS Biobank

To evaluate MIL model performance in a second external dataset, we used H&E sections of carotid plaques from the AtherOMICS biobank. AtherOMICS collects plaque tissue, peripheral blood, clinical and imaging data from patients undergoing endarterectomy surgery at LMU Klinikum, Munich, Germany. The study protocol has been published elsewhere^82^. AtherOMICS has received ethical approval from the Ethics Committee at the Faculty of Medicine of Ludwig-Maximilians-Universität (LMU) in Munich (registration nos. 121-09, 22-0135, 23-0772) and conforms to the Declaration of Helsinki guidelines. An overview of the cohort characteristics can be found in **Supplemental Table 15.**

Plaque samples of consenting study participants were collected during endarterectomy surgery in the operating room and processed immediately. Samples were divided into five-millimeter-thick sections. The section with the highest degree of atherosclerosis burden was fixed in 4% paraformaldehyde solution for 24h. After temporary storage in 70% ethanol, sections were decalcified in EDTA solution and subsequently embedded in paraffin. The paraffin blocks were then sent to the Core Facility Pathology and Tissue Analytics at Helmholtz München, Munich, Germany for histological processing.

Serial sections were collected in 1mm intervals throughout the sample and stained using H&E and Sirius Red for plaque morphology, as well as DAB-based anti-CD68 and anti-⍺SMA antibody stainings. Stained slides were imaged in brightfield and digitized on an Axioscan 7 scanner (Carl Zeiss, Oberkochen, Germany).

The resulting whole-slide .czi files of H&E-stained sections from 83 plaque samples were imported into QuPath (v.0.4.4),^83^ split into scenes containing one section each. Total plaque area and IPH were annotated as polygon overlays and labelled accordingly. IPH was identified using pre-defined annotation criteria.^82^ In addition, H&E sections from 16 unlabelled plaque samples were provided for this study.

### Multiple instance learning

#### WSI preprocessing

The main preprocessing steps applied to the WSIs are: tissue segmentation, tiling and feature extraction. By segmentation, the tissue is separated from the white background of the scanned image: to do this, a U-Net pre-trained on H&E stained samples from multiple tissue types was used. We subsequently retrieve from the tissue area squared tiles (512×512 pixels, corresponding to about 118×118 microns). Finally, features are extracted from these tiles using a ResNet50 convolutional neural network pre-trained on ImageNet.

#### IPH detection

The detection of intraplaque haemorrhage (IPH) from a stained whole slide image (WSI) relies on the additive multiple instance learning paradigm^84^.

Multiple Instance Learning (MIL) is a common framework in digital histopathology: tissue patches are grouped into bags, representing the full sample, and each bag is associated with a label. A bag is labelled positive if it contains at least one positive instance, and negative otherwise. Attention-based MIL is an extension that incorporates an attention mechanism aimed at assigning different levels of importance to instances within a bag. During training, the model learns attention weights, which enable it to focus on the most relevant instances while ignoring less significant ones. This mechanism helps in cases where the predictive information is localized in specific regions, providing also an attention map, which is useful for interpretability ^84^.

The Additive MIL model builds further upon attention-based MIL through a reformulation in the model’s calculations, by replacing attention weights with instance-wise contributions to the logits. The additive structure, beyond making the training more stable and robust, provides a clearer and direct interpretability of the tissue patches that actually contribute to the final output score.

All these improvements combined make the additive MIL model easier to validate, which helps to build trust in the model. Through the visualization of the attentive regions per class, a user can verify that the model’s predictions are based upon features that are supported by scientific evidence and similar to those identified by human experts ^84^.

The additive MIL model has been trained to predict IPH from the stained images based on the manual scoring (presence/absence); therefore, it performs binary classification, but it also provides patch-wise scores that can be considered to compute a proxy of IPH area.

#### IPH quantification

For each sample, the additive MIL model provides both the probability of the sample having IPH and the patch-wise scores that are used to compute that probability.

We first multiplied these scores by the sample size (to make them comparable across samples with different numbers of patches) and then applied the sigmoid function, to bring scores in the range [0,1]. Positive contributions (patches with a score > 0.75) are hypothesised to be predictive of the IPH area within the plaque, and their percentage with respect to the sample size (total number of patches) is the model-derived IPH area.

#### Manual Annotation and Model-Based Intraplaque Haemorrhage Quantification

To establish a reference standard for IPH detection, a clinical expert at UMCU manually annotated 95 WSIs, ensuring that all available histological stains were consistently labelled. The annotation process was performed using QuPath^83^, an open-source bioimage analysis tool (https://qupath.github.io/), allowing precise delineation of IPH regions. To prevent bias, the annotator was blinded to the model-generated results. These manually annotated regions were then used for evaluating the performance of our computational model in predicting IPH presence and spatial distribution.

For model-based IPH quantification, a pipeline was developed to generate patch-wise probability scores that indicate the likelihood of IPH in each region of a WSI. The model was applied to extracted patch features stored in .h5 files, along with the corresponding spatial coordinates. It returned logits representing the confidence of IPH presence, which were converted into probability scores using a sigmoid activation function. These scores were then scaled by the total number of extracted patches (bag size) to normalize predictions across different WSIs. A pseudo-mask was generated by selecting high-confidence patches, defined as those with a probability score exceeding 0.75, ensuring that only regions with a strong likelihood of IPH were retained.

To visually interpret model predictions, the identified positively contributing patches were overlaid onto the original slide, creating a greenish pseudo-mask representing the predicted IPH areas. Additionally, a spatial heatmap was generated by mapping patch-wise scores back to their corresponding tissue locations. The probability values were visualized using a ‘coolwarm’ colourmap, effectively highlighting regions with varying degrees of IPH presence. This heatmap was then superimposed onto a downscaled version of the WSI, allowing for a direct comparison between model predictions and manual annotations.

### SlideToolkit

The Athero-Express Biobank stores a large amount of scanned histological slides at high magnification. With the growing amount of whole slide images (WSIs), the demand for fast, accurate, and reproducible histological quantification increases. Since manual quantification is time-consuming and subject to observer variability, we developed slideToolKit^85^ to provide a toolset for automated feature analysis and histological quantification. It consists of a collection of open-source libraries and scripts that assist in processing a WSI at each step of the analysis procedure: acquisition, preparation, tiling, and analysis. For the acquisition phase, slideToolKit contains scripts to rename slides, convert slides into other TIFF formats when needed, or extract metadata (e.g. scan time, magnification, image compression). In the preparation step, directories for the downstream analysis are created and segmentation masks are retrieved, that have previously been made in the multiple instance learning (MIL) model’s WSI preprocessing step, to hide any artefacts in the WSI during the analysis step. The tiling step splits the WSI into smaller tiles used in the analysis step. The analysis step utilizes CellProfiler^86^ (version 4.2.6) to quantitatively measure histological features in the tiles.

SlideToolkit analyses a WSI patch by patch using a predetermined size. Because the Additive MIL models are trained using 512×512 patches, this resolution was also used in the SlideToolkit analysis. This also allowed us to extract the composition for the patches deemed to be in the IPH area. For each stain, we divided either the stained area or cell count by the total tissue size to accurately compare stain density between samples. For EVG, FIBRIN. GLYCC, and SR we used the staining area. We used the cell counts for CD34, CD68, CD66b, SMA, and HE.

We analysed the composition in two ways, first, we related the total composition between plaques with IPH versus plaques without IPH. Second, we compared the composition within plaques between IPH-positive and IPH-negative areas (determined from the attention heatmaps). Because each stain is from a serial section, the images for each stain do not always align with each other. Therefore, without image registration, we cannot use the H&E attention heatmaps to determine the IPH area for each stained image. Instead, we determine this IPH area for each stain using its own trained MIL model, which we use to determine the IPH area composition. As some trained models perform worse than H&E this might impact the composition results.

### Logistic regression ensemble model

Each histological stain binds to a specific biomarker, illuminating various cellular and extracellular components within tissue samples. For instance, EVG highlights elastin, Picrosirius Red emphasizes collagen, CD68 targets macrophages, and CD34 is specific for endothelial cells. Training models on individual stains allows for specialized feature extraction, enhancing the ability to identify features related to intraplaque haemorrhage (IPH) by focusing on the unique details each stain reveals.

To see what stain adds the most additional value, we used logistic regression ensemble models to combine H&E (the best-performing single stain model) with other stains to predict IPH presence. We combined the single stain model’s IPH probabilities of H&E and the other stain to create a set of 2 probabilities per sample. We used this to train a logistic regression model (Python 3.12.0, scikit-learn 1.4.1) to predict IPH presence. We trained using 10-fold cross-validation to validate the reliability and robustness of the logistic regression model performance.

### Prediction of reported symptoms

The prediction of symptoms from the integration of plaque phenotypes with electronic health records (35 features in total) was performed using a machine learning classifier (CatBoost).

In total 35 features were selected, including 23 clinical metadata variables: Age, Sex, Operated artery, Artery restenotic or de novo lesion, Preoperative degree of stenosis, History of MI, CAD and/or coronary intervention, Peripheral arterial occlusive disease, History of peripheral intervention, KDOQI classification, eGFR based on MDRD, LDL-cholesterol, HDL-cholesterol, Triglycerides, Systolic tension, Diastolic tension, BMI, Smoker status, Alcohol use, Diabetes status, Anti-coagulant use, Anti platelet drug use, Statin and/or LLD use. We also include 12 image-derived phenotypes: Neutrophil amount (rank normalized), Mast cell amount (rank normalized), Vessel density (rank normalized), Fat (binned at 40%), CD34 detected cells, CD68 detected cells, CD66b detected cells, SMA detected cells, FIBRIN detected area (ratio of plaque), EVG detected area (ratio of plaque), SR detected area (ratio of plaque), and the PHENOMICL’s quantified IPH area.

We fitted two models, one for stroke vs asymptomatic patients and a second for TIA vs asymptomatic patients. The two cohorts are composed of 1,080 (479 stroke, 601 asymptomatic) and 1,288 (687 TIA, 601 asymptomatic) patients respectively. CatBoost classifiers have been fitted for each task, using 10-fold cross-validation to measure performance; feature importances have been computed for each fold and then averaged.

### Differential gene expression analysis

Bulk RNA-Seq counts were transformed using variance stabilization transformation. We fitted linear models to perform differential gene expression on both continuous (amount of IPH, IHC markers quantification) and binary (presence of IPH) phenotypes. Specifically, we modelled gene expression as the sum of the phenotype of interest with other covariates (age, sex and sequencing batch id). The resulting p-values have been corrected using False Discovery Rate. Since the bulk RNA-Seq data contained both gene-level and transcript-level counts, we performed DGE at transcript resolution (without aggregation), where possible.

### Functional enrichment analysis and single-cell mapping

In order to identify biological processes, molecular functions, or pathways significantly associated with the set of differentially expressed genes, we performed functional enrichment analysis using decoupleR. We estimated transcription factor activity through the univariate linear model (providing *collectri* as network) and performed pathway activity inference with the multivariate linear model and the weights from *PROGENy*. In both cases, genes have been ordered by the absolute fold change from the DGE analysis. The Over Representation Analysis was run to infer enrichment scores of biological terms from the Human Molecular Signatures Database (MSigDB).

Lastly, as we had access to a single cell reference of atherosclerotic plaque and we wanted to identify the cell types involved in plaque vulnerability regulation, we used the *score_genes* function available in *Scanpy*, providing the significant (p-value < 0.05) DE genes as a gene list.

### Genome-wide association study

Genome-wide association study (GWAS) was performed using regenie (https://rgcgithub.github.io/regenie/)^87^, which consists of 2 steps. In the first step, a whole genome regression model is fitted using a subset of the genetic markers. This regression model captures a fraction of phenotype variance attributable to genetic effects. In the second step, the predictions from step 1 are used as covariates when each imputed SNP is tested.

It is important to filter out low-quality genetic data to improve the robustness of the analysis. First, using PLINK2 (https://www.cog-genomics.org/plink/2.0/)^88^, we converted the VCF files to PGEN (PLINK 2 binary) files to use in the regenie pipeline. We use hard-called genotypes in regenie step 1 and 2. Before processing with step 1 in regenie, the following quality control (QC) steps are taken and executed using PLINK2:

- Call Rate filtering: Removing variants with a call rate of less than 10%, as these may introduce noise and uncertainty.
- HWE filtering: Exclude variants with an HWE p-value higher than 1.0×10^−3^, as these might indicate genotyping errors or biases, making them unreliable.
- Minor Allele Frequency (MAF) filtering: Filter out variants with an MAF of less than 10%, as these could lead to spurious associations due to insufficient power.
- Pruning for independent variants: Remove variants with low linkage disequilibrium (LD) with an r^2^ threshold of 0.2. By pruning we select variants that provide unique information, improving the efficiency of the analysis.
- Exclude long-range LD blocks: Long-range LD can distort associations by creating artificial correlations. (**Supplemental Table 16**)
- Remove non-autosomal variants: Autosomal variants are generally present in both sexes and across different populations, making findings more universally applicable.
- Exclusion of A/T and C/G variants: Removing A/T and C/G variants helps minimize the impact of genotyping errors.

In both steps 1 and 2 of the regenie pipeline we used age, sex, principal component 1 (PC1), PC2, and year of surgery as covariates. Both regenie steps also used a blocksize (number of variants processed together (bsize)) of 1000.

Post-GWAS was done using Python (version 3.9.18) and GWASLab^89^ (version 3.4.44; https://cloufield.github.io/gwaslab/). During post-GWAS processing we perform GWASlab’s basic check function which makes sure that 1) SNPIDs are of the form chr:bp; 2) orders the data; 3 all alleles are capitalized; 4) does sanity checks on data. After this we remove duplicate and multi-allelic variants and harmonize the data with Human Genome Build 38 to 1) make sure alleles are oriented according to the reference; 2) assign rsIDs; 3) flip alleles (and effect size) when necessary. Next, we check for palindromic variants and indistinguishable indels and remove these. Last, we filter based on the Delta Allele Frequency (DAF) in GWASLab, which is calculated as the difference between the effect allele frequency (EAF) in the summary statistics and the alternate allele frequency in the reference VCF/BCF file (>-0.12 and <0.12). This step aims to exclude variants with large discrepancies in allele frequencies between the summary statistics and the reference VCF/BCF file.

### Spatial transcriptomic analysis

For the spatial transcriptomics analyses we used the 10X Genomics Visium HD technology. The HD slide contains a continuous array of oligonucleotides arranged into about 11 million 2 x 2 µm barcoded squares without gaps, within a 6.5 x 6.5 mm capture area, preserving the original location of mRNA in the tissue. We selected 8 samples from the Athero-Express Biobank based on overlap with the 46 single-cell RNAseq samples, availability of paraffin embedded tissue, whole-slide images of all stains (H&E, EVG, Fibrin, Glycophorin C, CD34, CD66b, CD68, SMA, picrosirius red), and RNA quality of FFPE samples. The 8 samples were age (within 10 years) and sex matched (**Supplemental Table 17; Supplemental Figure 12**).

#### Sample processing and RNA quality assessment

RNA quality of FFPE carotid artery samples was evaluated using DV200 > 30% - the average DV200 was 57.1% for the 8 selected samples. Sections were cut to 5 μm thickness as per the Visium HD FFPE Tissue Preparation Handbook (CG000684, 10x Genomics), mounted on TrueBond 380 slides, and stored at -80°C. Slides were used within 1 week to prevent RNA degradation.

Tissue sections were deparaffinized, stained with H&E, and imaged at 20x magnification using a Leica Thunder Imager (Leica). After imaging, the coverslip was removed, and sections were destained and permeabilized with 1% SDS, then incubated for 1 hour in 70% cold methanol, and decrosslinked.

#### Visium HD library preparation

Samples were processed following the Visium CytAssist Spatial Gene Expression Reagent Kits User Guide (CG000685), with hybridization performed at 50°C for 17 hours, followed by washes and probe ligation. The transcriptomic probes were transferred onto the Visium HD slide using the CytAssist instrument for 30 minutes at 37°C to capture the probes while preserving their spatial localization. The HD slide features a continuous lawn of oligonucleotides arranged into approximately 11 million 2 x 2 µm barcoded squares without gaps, within a 6.5 x 6.5 mm capture area, thereby retaining information about the mRNA’s original location in the tissue. After releasing the probes from the HD slide, the captured mRNA molecules were reverse transcribed into cDNA and tagged with a spatial barcode to create libraries. These libraries were indexed with the Dual Index Kit TS Set A (PN-1000251, 10x Genomics) and purified using SPRIselect (Beckman Coulter). Each of the 4 sample pairs was processed across two capture areas (positions A1 and D1), yielding 8 libraries (tags 1A/1B– 4A/4B) with an average fragment size of 313–341 bp and library concentrations of 47.5–234.8 nM prior to pooling.

#### Novogene

After library preparation, samples were sent for sequencing to Novogene (flowcell lane 22GKHGLT4_L8; 150 bp paired-end reads). Sequencing yielded an average of 540.9 million raw reads per library (range 489.3–574.1 million; 81.1 Gb average raw data per library; 4,326,902,158 reads total across the 8 libraries), with an average sequencing error rate of 0.06%, GC content of 45.8% (range 45.3–46.6%), Q20 of 77.1%, and Q30 of 61.2%.

#### Spatial data quality control

Reads and images were processed using `SpaceRanger` version 4.0.1(10x Genomics, https://www.10xgenomics.com/support/software/space-ranger/latest), reads were aligned to the Probe Set v2 (GRCh38 2020-A) and to corresponding bins, which were mapped back to the Cytassist and H&E-stained tissue images. The filtered count matrix was used for downstream processing and data analysis.

To account for low sequencing coverage due to the sample complexity, Spaceranger was run with 8, 16, 18, 20, 22, 24, 26, 28 and 32 --custom-bin-size settings and a comprehensive QC and optimization of filtering parameters was performed subsequently.

Mitochondrial genes were removed for all analyzed samples. Multiple filtering thresholds were tested across four filtering parameters. Bins were filtered based on the minimum number of detected genes (1, 3, 10 or 50) and a minimum number of total transcript counts (1, 3, 10 or 50). For genes, a requirement of being detected in a minimum number of bins (1, 3, 10 or 50) and to have a minimum number of transcripts (1, 3, 10 or 50) was introduced, resulting in 256 quality-control tested settings. For each of the tested parameter combinations, low quality spatial bins and low abundant genes were removed. Filtered data were log transformed with the `pp.log1p` function from `scanpy` (v1.11.0). Principal component dimensionality reduction was performed with `scanpy pp.PCA` function using 20 components, and a nearest-neighbor graph was constructed from the PCA space and then leiden clustering was performed with different resolutions (from 0.1 to 1.0 were tested in increments of 0.1) and `flavor = “igraph”` using `tl.leiden` function from `scanpy`. Resolution testing was stopped early if the number of clusters exceeded 55. The number of kept genes, bins as well as number of identified clusters were recorded.

Next, from the generated QC parameter space, parameter combinations resulting in fewer than 50 Leiden clusters and retaining more than 50% of spatial bins were selected for further testing. A Pareto-based optimizer was introduced to identify parameter combinations that minimized the bin size and the number of Leiden clusters generated (better data grouping), while maximizing the slide spatial bin coverage. Parameter combinations were ordered by increasing bin size, increasing cluster number, and decreasing coverage. For each of the analyzed bin sizes, combinations that provided an improvement in coverage were retained as Pareto-optimal solutions.

Upon visual inspection of the number of genes expressed per bin and read per bin on VisiumHD slides, we selected the optimal parameter: bin size of 24um, and filtered the data using scanpy pp.filter_genes (min_cells=3, min_counts=3) and scanpy pp.filter_cells (min_genes = 1, min_counts = 1).

#### Spatial gene expression analysis

Gene expression of the top 10 upregulated genes from the model-IPH DGE were spatially plotted. Data from the 10 genes were extracted, filtered on the 99th percentile (to remove extreme outliers), and min-max normalized. In addition, cell type specific gene lists for level 1 (L1) labels were derived from our single-cell RNA sequencing data through MetaPlaq v2^53^.

### Differential gene expression

Spatial RNA-Seq DGE was performed between IPH and non-IPH regions based on the model’s heatmaps. The spatial data and IPH heatmaps data have been manually aligned to overlap spatial bins and IPH tiles. The spatial gene counts were summed per region (IPH or non-IPH) to create a pseudobulk RNA set. This resulted in 16 pseudobulk RNA sets (8 IPH, 8 non-IPH) to be used for the differential expression analysis. One sample was removed due to low gene counts (< 15,000 total counts) in the IPH pseudobulk RNA set, likely reflecting a small IPH area. Differential gene expression analysis was performed using the DESeq2 (version 1.50.2) library in R (version 4.5.2). Genes with a low gene count (less than 10 counts across all remaining samples) were also filtered out. The resulting p-values have been FDR (False Discovery Rate) corrected.

### Imputation

For the survival analysis using a Cox Proportional Hazards Survival Regression model, missing values in the clinical data were imputed using the MICE^90^ package in R (version 4.4.1). This package helps to impute missing values with plausible data which are drawn from a distribution specifically designed for each missing data point.

Missing data can be classified into three categories: missing completely at random (MCAR), missing at random (MAR), and missing not at random (MNAR). MCAR means that the probability of missing a data point is the same for all samples. This implies that the cause of the missing data is unrelated to the data. MAR is broader than MCAR. It means that the probability of missing a data point is the same within groups defined by the observed data. MNAR means that the probability of missing a data point depends on unknown reasons. For example, some patient measurements are not taken if the patient’s age is below 40. MNAR is the most complex imputation case and requires varying strategies^91^.

In our models features like CRP and HDL, used by Hellings *et al*, are missing in more than 40% of the patients (**Supplemental Figure 6)**. For all features with missing values, we assume a case of MCAR or MAR. For all missing variables, we used the predictive mean matching (PMM) imputation method. It is an easy-to-use, versatile, and robust method. PMM fills in a missing entry by randomly picking the value from a set of 5 candidate samples. The set of candidate samples is determined by looking at which regression-predicted values are closest to the regression-predicted value for the missing value from the simulated regression model. This method makes sure that all values are plausible and fall within the observed data range.

Using multiple imputation we create multiple versions of a complete dataset. Each analysis starts with the incomplete dataset and replaces the missing data with plausible data values, based on a determined imputation method. After creating multiple versions of the dataset, they are pooled together into a single final estimate. The differences between imputed values across these datasets reflect the uncertainty of the imputation. We ran the imputation for 10 iterations with 100 imputed datasets per iteration. The average certainty of the imputation is 0.903, with a more stratified certainty per variable in **Supplemental Table 10**. For continuous variables, the certainty measure is based on the standard deviation ratio between imputed and observed values. For categorical variables, the certainty is based on the mode proportion of imputed values compared to observed values. Mode proportion determines how well the dominant category in imputed values matches the observed values. This certainty measure ranges from 0 to 1, where < 0.5 is a very low certainty (Imputed values differ significantly from the original distribution) and > 0.9 is a high certainty (Imputed values closely match the original distribution)

### Cox Proportional Hazards Survival Regression

To validate the model’s results, we replicated results from studies by Hellings *et al*.^63^ and Vrijenhoek *et al.*^34^. The study by Hellings *et al.* showed that intraplaque haemorrhage, scored as Yes/No, was significantly correlated with the event rate after surgery (HR=1.7, 95% CI, 1.2– 2.6). The subsequent study by Vrijenhoek *et al.* stratified their patients by sex and found that IPH was only significantly associated with the event rate in men (HR=1.9, 95% CI, 1.2–2.8). In women, there was no significant correlation (HR=1.0, 95% CI, 0.6–1.7).

To reproduce these results we created a Cox proportional hazards survival regression in Python (version 3.12.0) using the package “lifelines”. We used the same covariates as used by Hellings *et al.* While they used the same Athero-Express dataset, exact variable names were not provided. We chose all variables based on the description given in the paper (**Supplemental Table 12**). The study by Hellings *et al.* only included patients up until March 11, 2008, which we therefore also used as a cutoff.

### Cell-cell Interaction Analysis

Cell-cell communication analysis was performed using the MetaPlaq v2 atlas^53^. Cell-cell communication analyses were performed in CellChat^92^ (version 2) using the default workflow and granular cellular subtype, ‘level2 annotations’, were used for cell-type annotation. The normalized count matrices were used for all analyses as recommended by the authors. To compare cell-cell interactions across lesion stages (advanced vs early), comparison analysis was accomplished in CellChat. Significant interactions altered across lesion stage were determined using thresholds recommended by the authors (p.val < 0.05, | LFC | > 0.05).

## Data availability

Athero-Express Biobank Study: https://doi.org/10.34894/4IKE3T

GWAS summary statistics: GWAS Catalog accession identifiers GCST90559271 and GCST90559272

DGEA results: https://doi.org/10.34894/ZODL42.

## Code availability

Model: https://github.com/CirculatoryHealth/PHENOMICL

Downstream analyses: https://github.com/CirculatoryHealth/PHENOMICL_downstream

GWASToolKit: https://github.com/swvanderlaan/GWASToolKit

slideToolKit: https://github.com/swvanderlaan/slideToolKit

## Links

Roche Ventana iScan HT: https://diagnostics.roche.com/global/en/products/instruments/ventana-iscan-ht.html Hamamatsu C12000-22 Digital slide scanner: https://www.hamamatsu.com/eu/en/product/life-science-and-medical-systems/digital-slide-scanner/index.html

1000G high coverage phase 3 data b38: https://ftp.1000genomes.ebi.ac.uk/vol1/ftp/data_collections/1000G_2504_high_coverage/working/20220422_3202_phased_SNV_INDEL_SV/

## Acknowledgements

We are thankful for the support of the Netherlands CardioVascular Research Initiative of the Netherlands Heart Foundation (CVON 2011/B019 and CVON 2017-20: Generating the best evidence-based pharmaceutical targets for atherosclerosis [GENIUS I&II]), the ERA-CVD program ‘druggable-MI-targets’ (grant number: 01KL1802), the Leducq Foundation ‘PlaqOmics’, ‘AtheroGen’ and ‘COMET’, and the Chan Zuckerberg Initiative ‘MetaPlaq’. The research for this contribution was made possible by the AI for Health working group of the EWUU alliance (https://aiforhealth.ewuu.nl/). The collaborative project ‘Getting the Perfect Image’ was co-financed through use of PPP Allowance awarded by Health∼Holland, Top Sector Life Sciences & Health, to stimulate public-private partnerships.

We would like to thank all the (former) employees involved in the Athero-Express Biobank Study of the Departments of Surgery of the St. Antonius Hospital Nieuwegein and University Medical Center Utrecht for their continuing work. In particular we would like to thank (in no particular order) Marijke Linschoten, Arjan Samani, Tim Bezemer, Tim van de Kerkhof, Joyce Vrijenhoek, Ben van Middelaar, Sander Reukema, Sander M. van de Weg, Arjan H. Schoneveld, Robin Reijers, Joëlle van Bennekom, and Bas Nelissen for their work and support on ExpressScan and slideToolKit. Lastly, we would like to thank all participants of the Athero-Express Biobank Study; without you these kinds of studies would not be possible.

## Contributions Funding sources

SWvdL is funded through EU H2020 TO_AITION (grant number: 848146), and EU HORIZON MIRACLE (grant number: 101115381). TSP and SWvdL are funded through HealthHolland PPP Allowance ‘Getting the Perfect Image’. CLM is funded by National Institutes of Health (NIH) grants (R01HL148239 and R01HL164577), Leducq Foundation network grant ‘COMET’ (24CVD02), and American Heart Association Transformational Project Award (24TPA1300556). CLM and SWvdL are funded through EU HORIZON NextGen grant (101136962), NIH grant U01DK142283, and Chan Zuckerberg Initiative Data Insights grant ‘MetaPlaq’. AtherOMICS has received funding by the German Research Foundation (DFG; as part of the Emmy Noether programme [GZ: GE 3461/2-1, ID 512461526] to MKG; the Munich Cluster for Systems Neurology [EXC 2145 SyNergy, ID 390857198] to MKG; and the Collaborative Research Center 1744 [ID 548585053] to MKG), the Fritz-Thyssen Foundation (grant ref. 10.22.2.024MN to MKG), the Hertie Foundation (Hertie Network of Excellence in Clinical Neuroscience, [ID P1230035] to MKG), and the LMU-TAU Research Cooperation Program (to MKG), as part of the Excellence Initiative of the German federal and state governments. HeCES2 is funded by State Research Funding (VTR) awarded by the Ministry of Social Affairs and Health, Finland (TYH2024401), EU HORIZON NextGen grant (101136962).

## Journal subject terms

Atherosclerosis, Intraplaque haemorrhage, cardiovascular disease, multiple instance learning, machine learning, genetics

