## Supplementary Material for "Intraplaque haemorrhage quantification and molecular characterisation using attention-based multiple instance learning"

*Accompanying*

*by*

*Francesco Cisternino, Yipei Song, Tim S. Peters, Maria Murach, Patrick Hart, Jose Verdezoto Mosquera, Laura Mäkitie, Mikko I. Mäyränpää, Luka Zivkovic, Roya Batool, Julian Louma, Abdalla Tarek Marei, Nikolaos Tsilimparis, Ana Karina de Oliveira, Roderick Westerman, Gert J. de Borst, Ernest Diez Benavente, Noortje van den Dungen, Petra Homoed-van der Kraak, Dominique P.V. de Kleijn, Joost Mekke, Michal Mokry, Gerard Pasterkamp, Hester M. den Ruijter, Evelyn Velema, Petra Iljäs, Marios K. Georgakis, Clint L. Miller, Craig A. Glastonbury, S.W. van der Laan.*

### Index of content

**Index of content** **1**
**Supplemental Figures** **2**
**Supplemental Tables** **12**

### Supplemental Figures

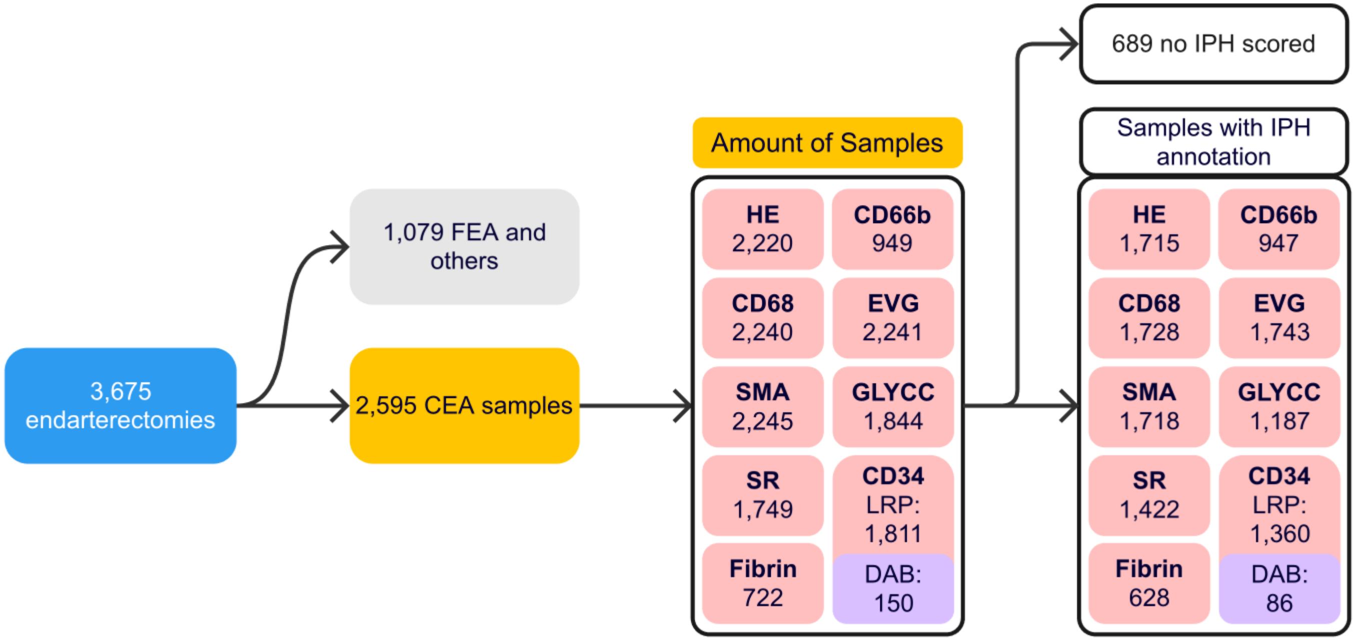


**Supplemental Figure 1 Patients included.** As of 09-08-2023 the AtheroExpress included 3,675 patients undergoing arterial endarterectomy, of which 2,595 underwent carotid endarterectomy (CEA). Not all staining types are available for all samples, nor is IPH scored in all samples. Of the 2,595 CEA samples, 689 samples are not scored for IPH. However, the number of IPH-scored samples differs across stains because each stain is available for only a subset of samples (i.e. within a given stain-specific subset, the number of not scored IPH samples is lower than 689). For example, among 2,220 H&E samples, only 505 are not scored for IPH. FEA: femoral endarterectomy (includes femoral and iliac arterial plaques).

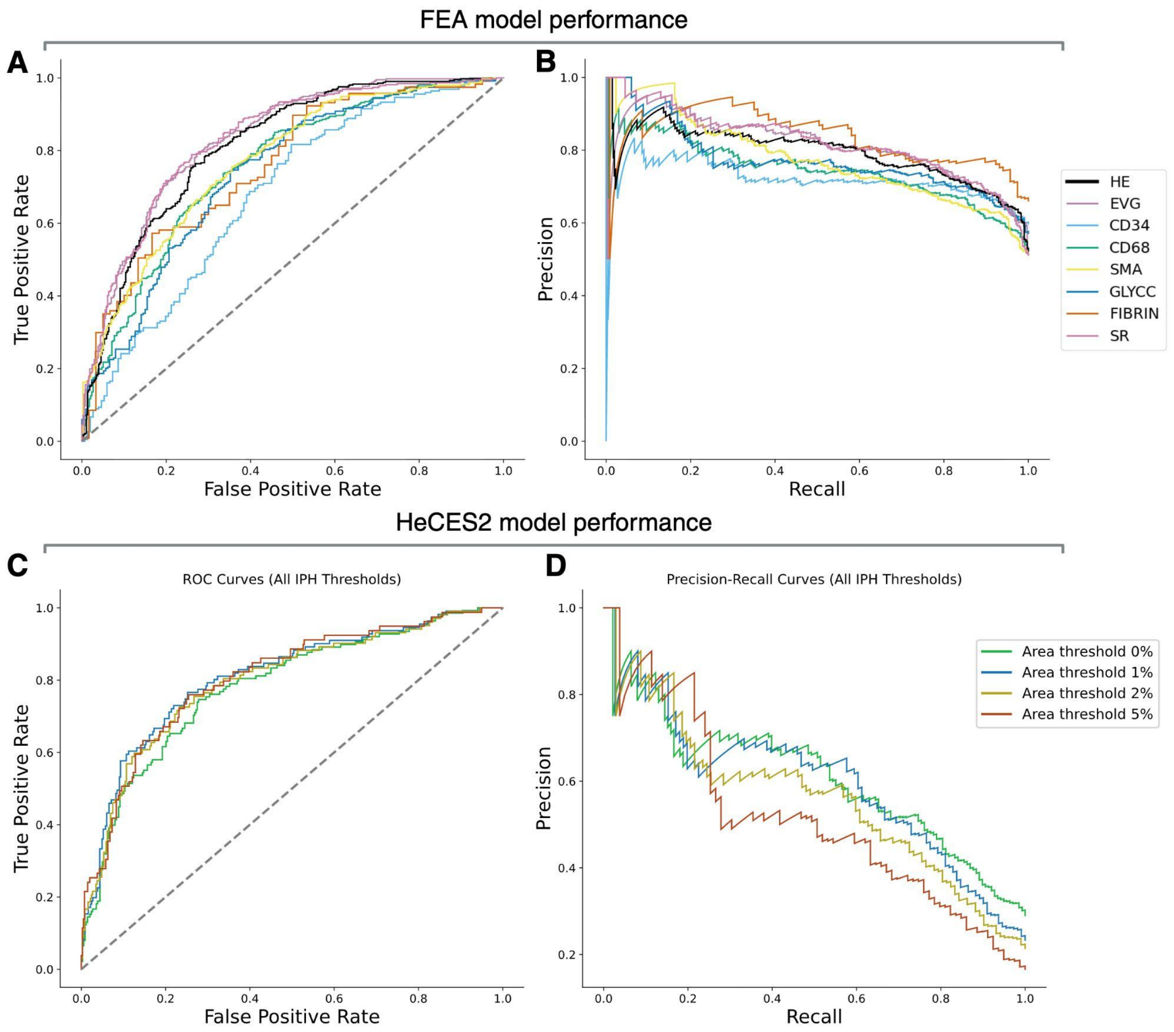


**Supplemental Figure 2: Additive MIL model performance on femoral endarterectomy (FEA) and HeCES2 validation cohorts.** Receiver Operating Characteristic (ROC) curve (**A, C**) and Precision-Recall (PR) curve (**B, D**) for IPH classification model validated on the FEA cohort (**A, B**) and HeCES2 cohort (**C, D**). The FEA cohort panels (**A, B**) show performance from models trained on stains: H&E, Elastin Verhoeff–Van Gieson (EVG), CD34, CD68, Smooth Muscle Actin (SMA), glycophorin C (GLYCC), Fibrin, and Sirius Red (SR). EVG and SR achieved a similar high AUC performance (AUROC = 0.832 and 0.831 respectively), outperforming H&E (AUROC = 0.813). The HeCES2 cohort panels (**C, D**) show H&E model performance at different thresholds of the manually scored IPH area (e.g. a threshold of 2% classifies manual-IPH area > 2% as ‘IPH yes’, otherwise <= 2% as ‘IPH no’).

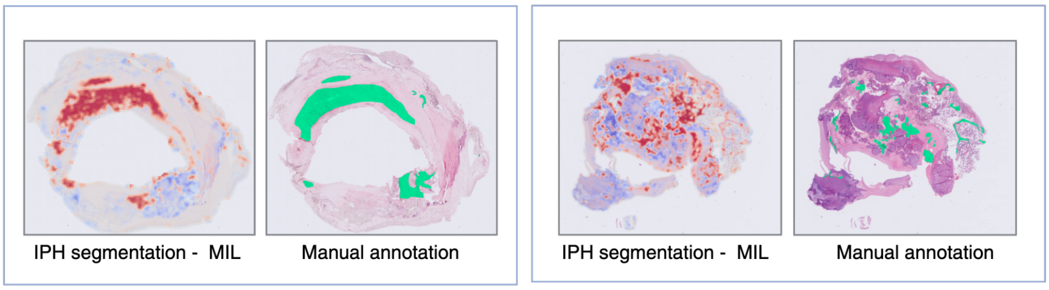


**Supplemental Figure 3: Additive MIL attention heatmaps**. MIL-derived segmentation of IPH validated with manual annotations in the Athero-Express cohort.

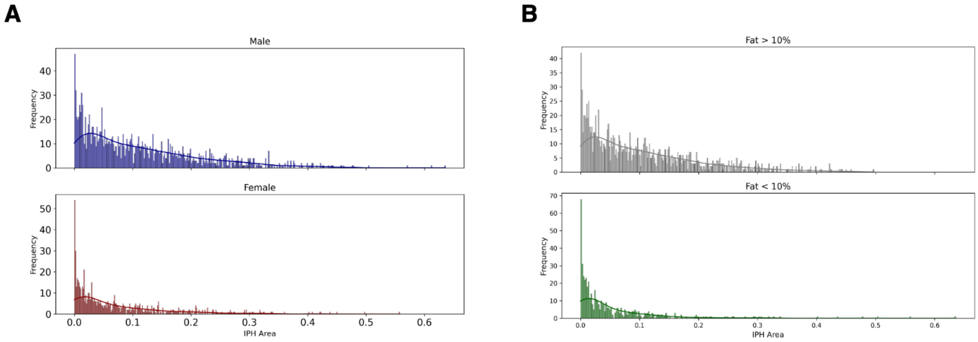


**Supplemental Figure 4: Model-IPH abundance distribution across sex and fat content**. Histogram showing differences of model-IPH abundance distribution between male and female (**A**) and between samples with > 10% and < 10% fat content (**B**).

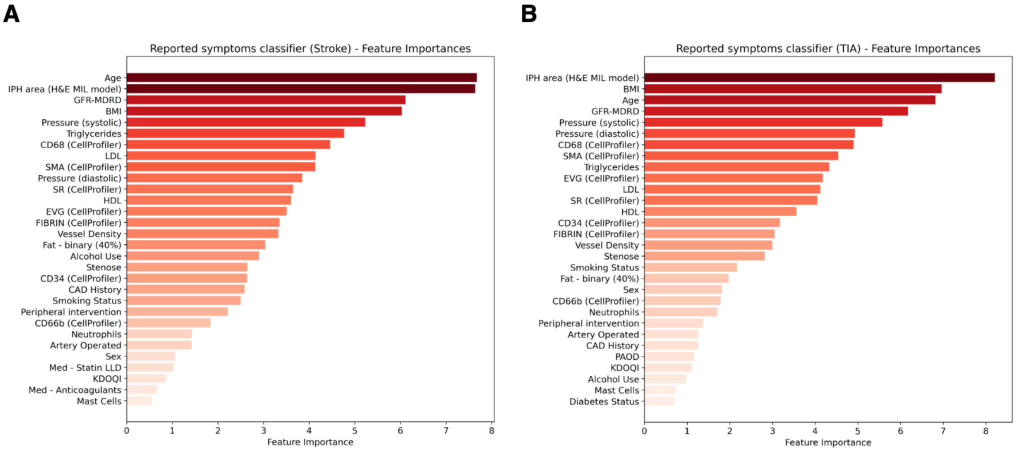


**Supplemental Figure 5: Symptoms classifier feature importance**. Ranked feature importance from CatBoost classifier for the prediction of stroke (**A**) and transient ischemic attack (**B**) occurrence from plaque morphological phenotypes and electronic health records. (BMI: Body Mass Index; GFR-MDRD: Glomerular Filtration Rate estimated using the MDRD equation; LDL: Low-Density Lipoprotein; HDL: High-Density Lipoprotein; CAD: Coronary Artery Disease; PAOD: Peripheral Arterial Occlusive Disease; KDOQI: Kidney Dialysis Outcomes Quality Initiative)

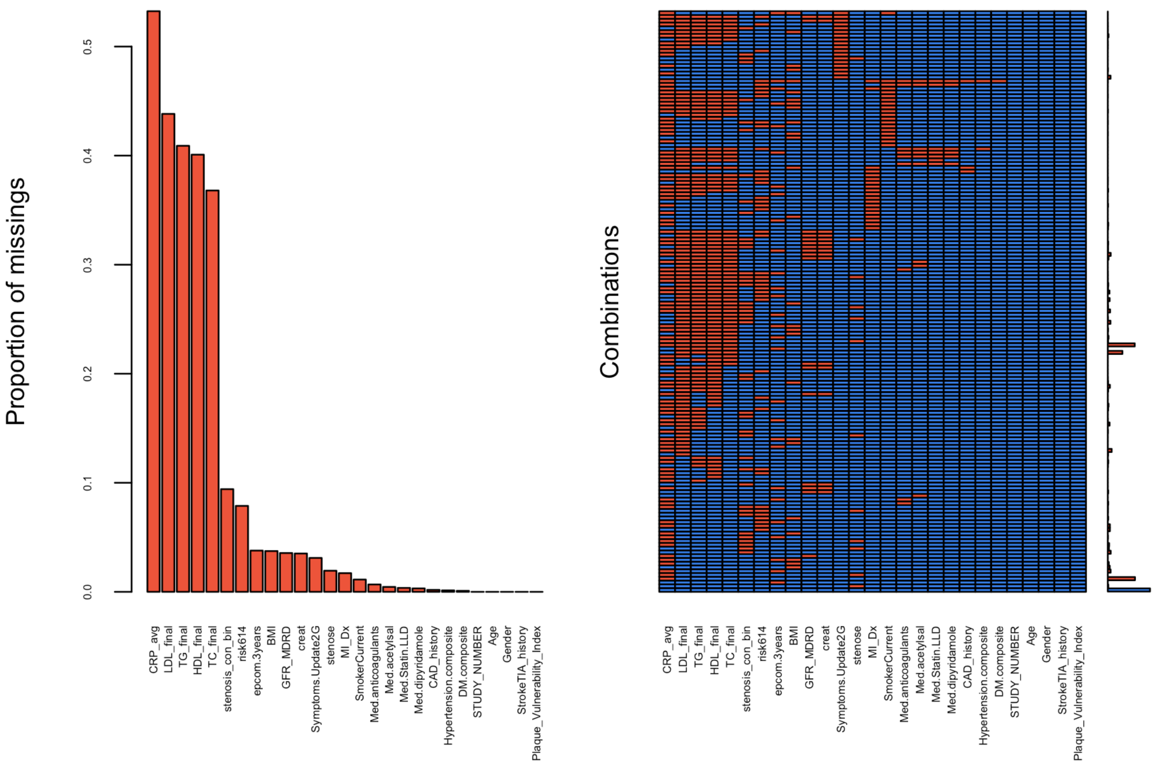


**Supplemental Figure 6: Missing clinical data across the cohort.** Histogram of missing clinical data (x-axis showing variables) (**Left**). Pattern of missing data, y-axis showing different combinations of missingness and x-axis showing the clinical data variables (**Right**). Histogram showing the number of samples per missingness combination (**Right-most**).

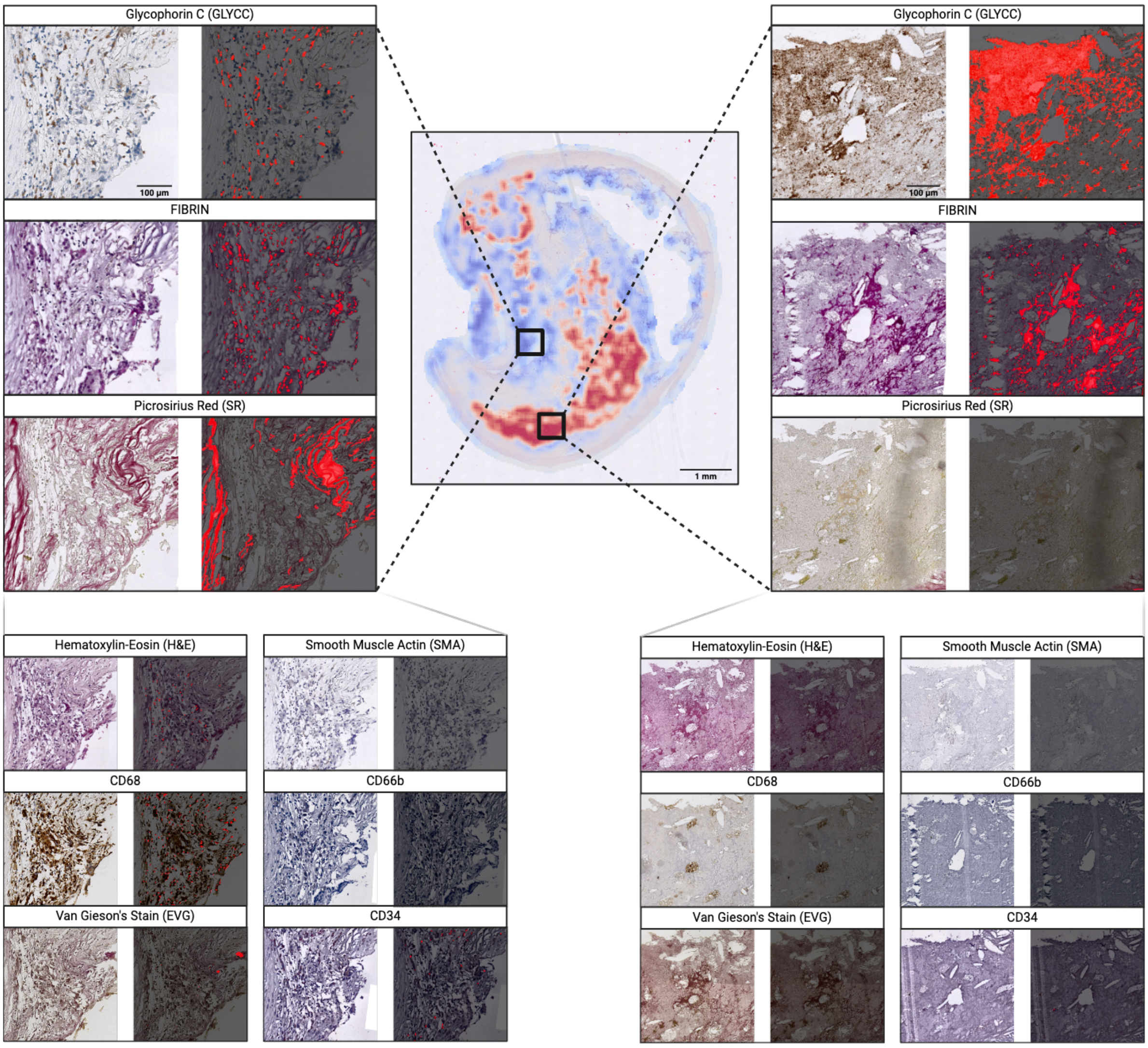


**Supplemental Figure 7**: **CellProfiler differential composition overview.** Plaque composition comparison between IPH positive vs. negative area. GLYCC, FIBRIN, and SR stains show the most significant difference. The red overlay (dark right-placed images) specifies the stained area as determined by slideToolKit[10,35] and CellProfiler[9] (completely dark indicates little to no staining). (**Left**) enlarged stain patches of IPH negative area. (**Right**) enlarged stain patches of IPH positive area. (Smaller bottom images are scaled-down versions of the larger top images, with the same scale bar applied proportionally.)

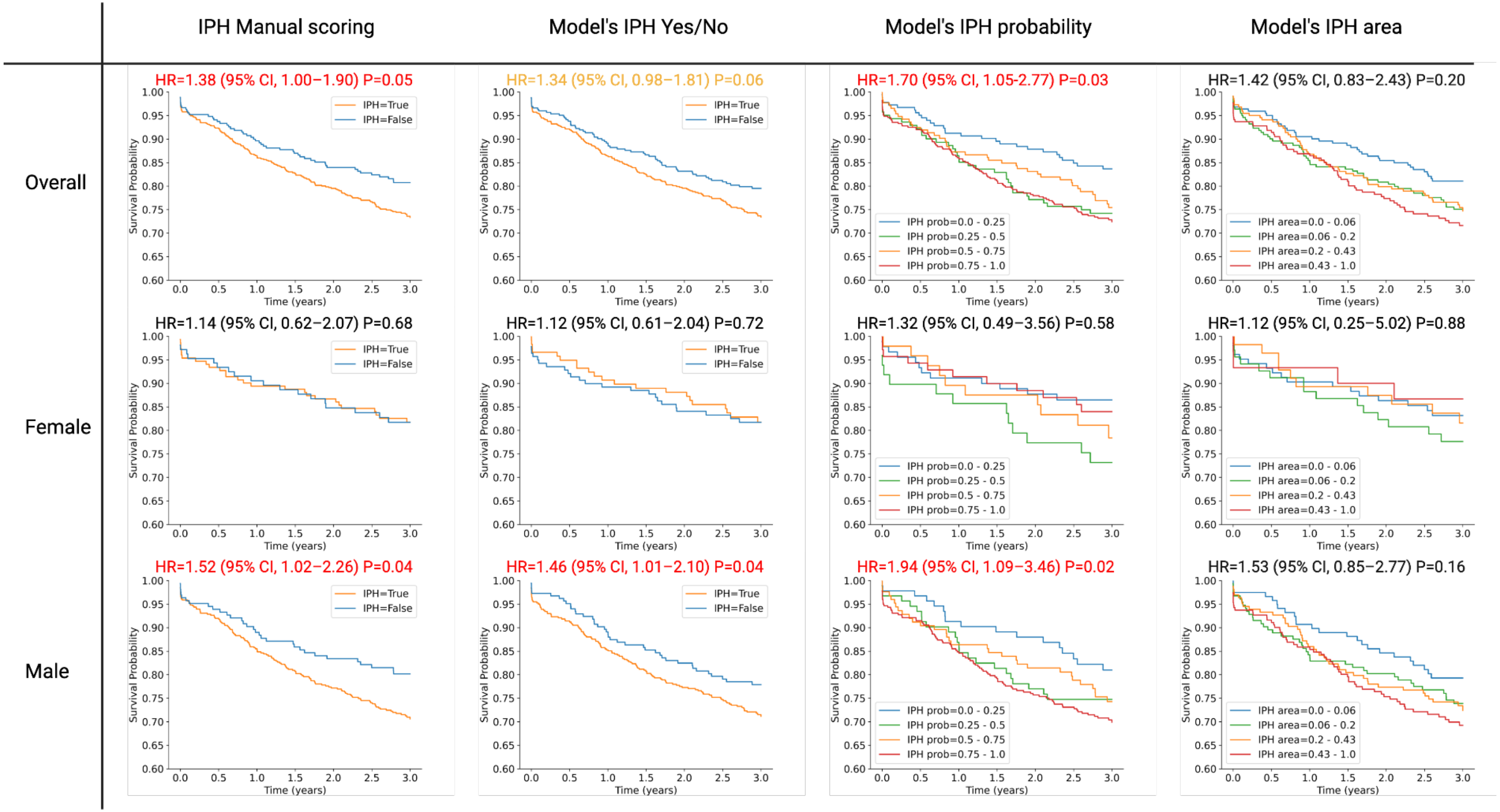


**Supplemental Figure 8**: **Cox regression Kaplan-Meier curve of manual and predicted IPH scores.** Showing how changes in a covariate affect the survival probability. Showing imputed data before March 11, 2008.

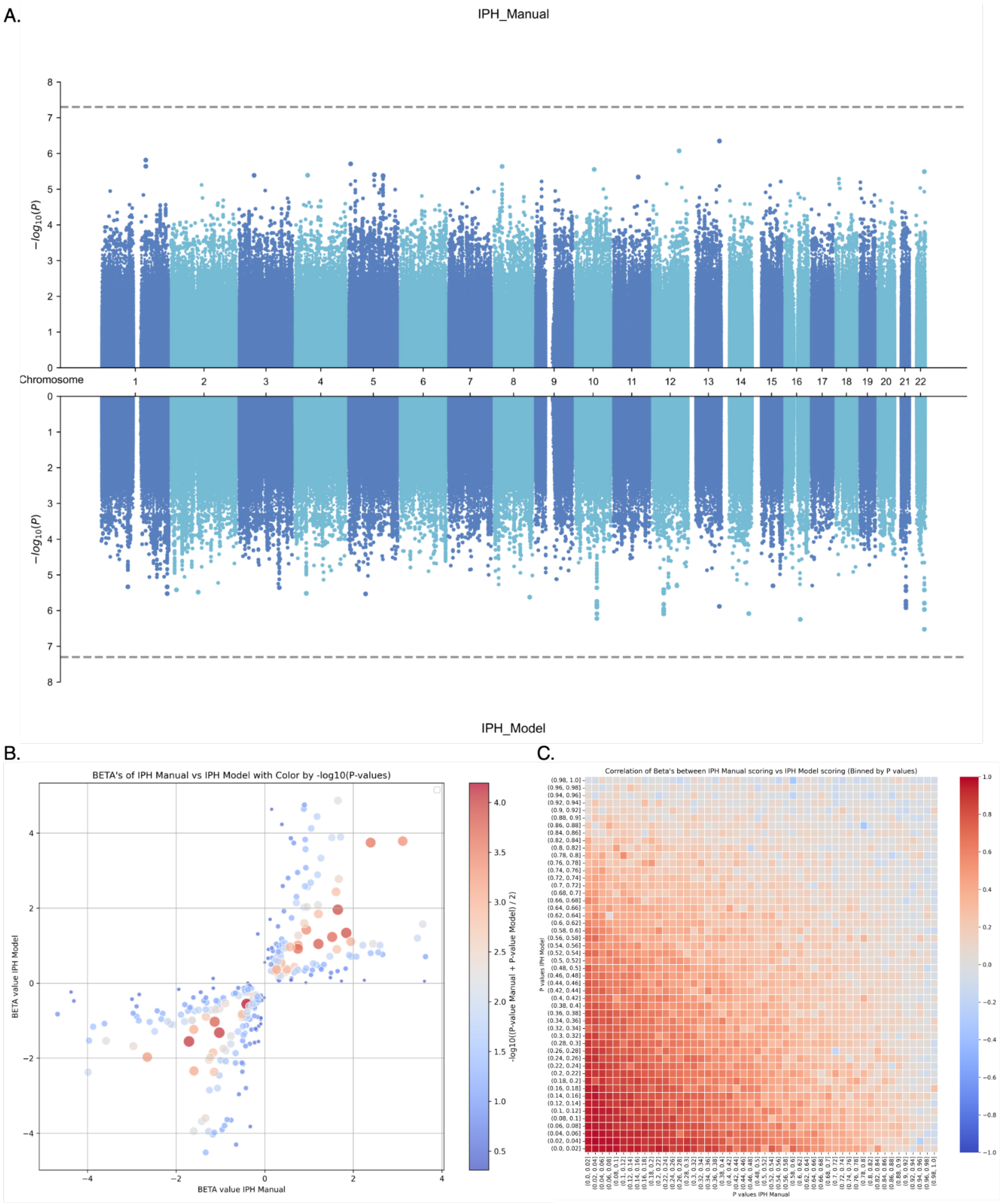


**Supplemental Figure 9: Genome-Wide Association Study between manual and predicted IPH scoring.** MIAMI plot of GWAS between manual and predicted IPH scoring (**A**). Scatter plot of genes with the lowest p-value between manual and predicted IPH scoring, axes show the beta values of the two GWAS's (**B**). Correlation plot of beta values binned by p-value between manual and predicted IPH scoring (**C**).

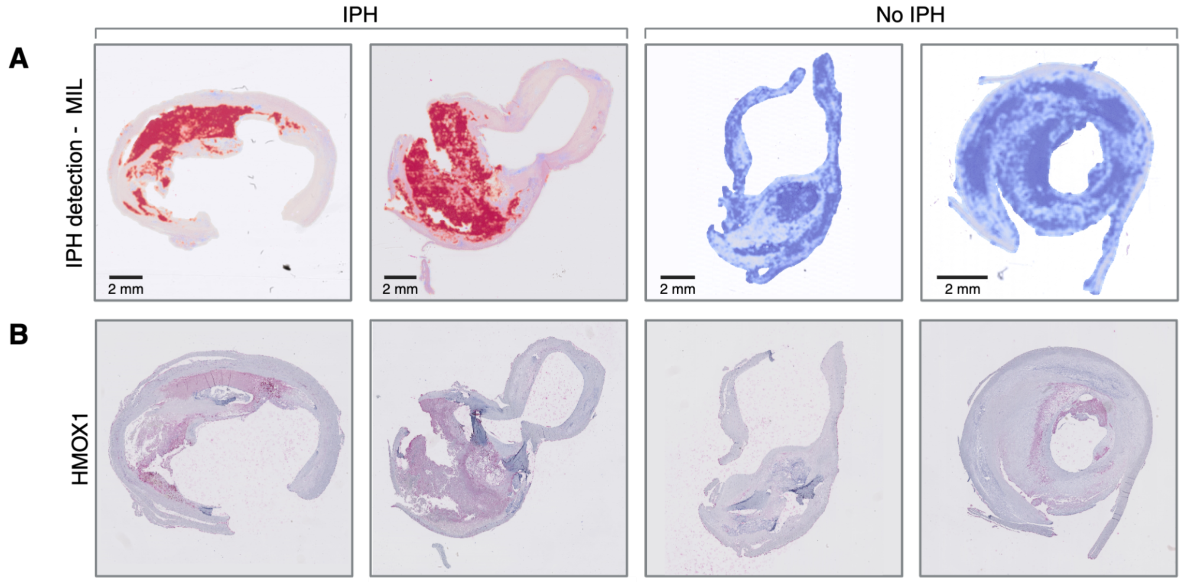


**Supplemental Figure 10: HMOX1 staining correlates with model-IPH area**. Intraplaque haemorrhage localization (**A**) from H&E images compared to HMOX1 IHC staining (**B**), showing that areas of IPH express more HMOX1 staining.

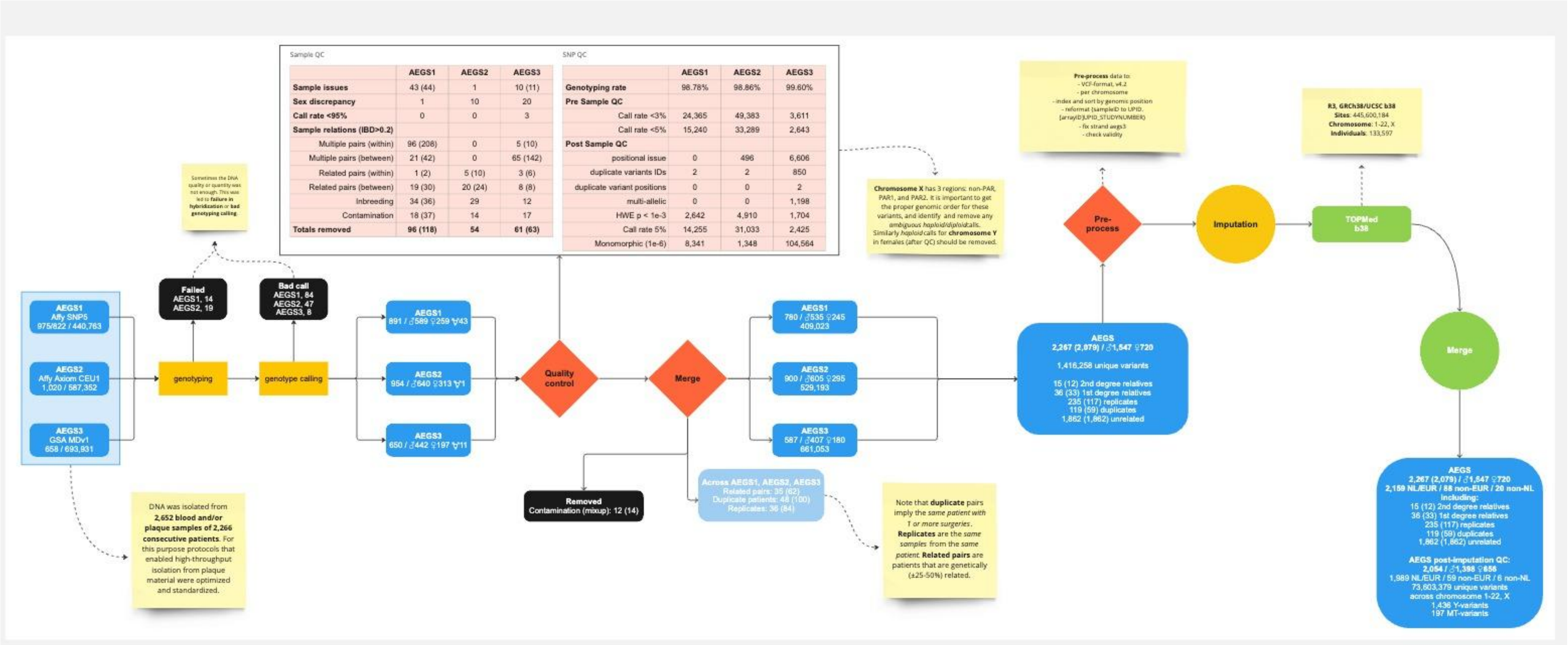


**Supplemental Figure 11: Full quality control and assurance workflow of the Athero-Express Genomics Studies.** AEGS1: Athero-Express Genomics Study 1 genotyped with Affymetrix SNP 5.0. AEGS2: Athero-Express Genomics Study 2 genotyped with Affymetrix Axiom CEU. AEGS3: Athero-Express Genomics Study 3 genotyped with Illumina GSA MD v1. These represent three separate but sequential experiments executed using samples from the Athero-Express Biobank Study (van der Laan et al. 2015; van der Laan et al. 2018). Note that patients in the Athero-Express can be operated on multiple arteries (for example, left and right carotid), thus some patients have multiple different samples - from a different arterial bed - available. Thus there are duplicate (or multiplicate) samples, derived from the same patients but different arterial beds. There are also replicate samples, derived from the same patient and same arterial bed.

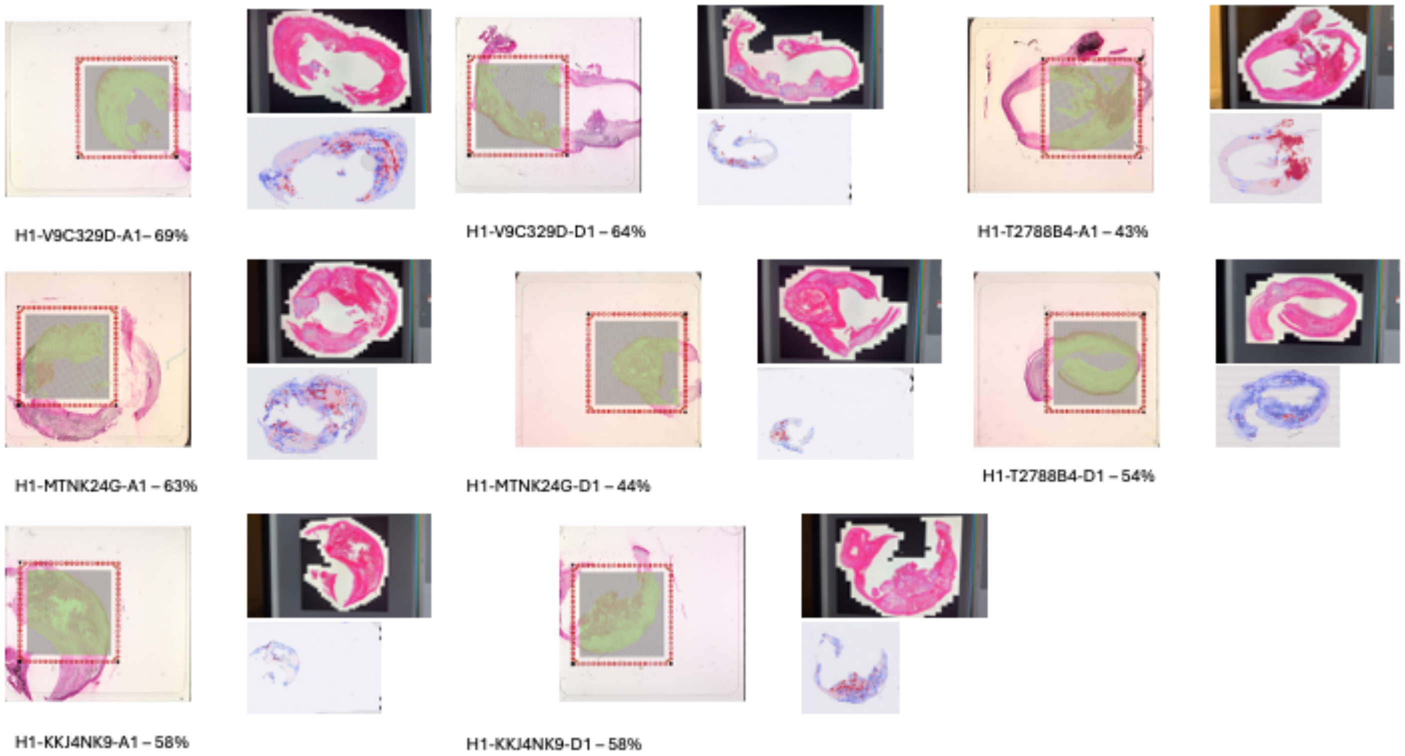


**Supplemental Figure 12: Visium HD histology imagery.** For each of the 8 samples three thumbnail-size images are given. The large image (left side) shows the Capture Area used. The top right image shows the H&E stain made per Visium HD protocol as reference. The bottom right image shows the localization of the predicted IPH area. The percentage DV200 is indicated for each sample.

### Supplemental Tables

**Supplemental Table 1: Baseline characteristics of the Athero-Express Biobank Study.** Intraplaque haemorrhage reported is manually scored. Including percentage missingness.

| Characteristic | All patients (n=2,595) | Male (n=1,803) | Female (n=792) | p-value | Missing (%) |
|---|---|---|---|---|---|
| Age (y (SE)) | 69.24 (9.22) | 69.21 (8.93) | 69.32 (9.85) | 0.773 | 0.0 |
| BMI (kg/m2 (SE)) | 26.525 (4.106) | 26.545 (3.549) | 26.480 (5.171) | 0.718 | 3.7 |
| Current Smoker (% (n)) | 35.2 (873) | 32.8 (568) | 40.8 (305) | <0.001 | 4.5 |
| Diabetes mellitus (% (n)) | 23.4 (607) | 23.4 (607) | 21.6 (171) | 0.176 | 0.1 |
| Hypertension (% (n)) | 85.1 (2207) | 84.6 (1526) | 86.3 (681) | 0.297 | 0.1 |
| Hypertensive drugs (% (n)) | 76.3 (1972) | 76.4 (1374) | 76.3 (598) | 0.996 | 0.5 |
| Anticoagulants (% (n)) | 11.2 (288) | 12.5 (224) | 8.2 (64) | 0.002 | 1.0 |
| Use of statin and/or LLD (% (n)) | 80.2 (2071) | 80.0 (1438) | 80.7 (633) | 0.694 | 0.5 |
| Glomerular filtration rate (mL/min (SE)) | 73.81 (21.88) | 74.84 (21.76) | 71.44 (22.01) | <0.001 | 4.2 |
| Restenosis (% (n)) | 5.1 ( 130) | 4.0 (71) | 7.6 (59) | | 1.4 |
| Symptoms (% (n)) | | | | 0.039 | 0.5 |
| TIA | 39.4 (1017) | 38.5 (692) | 41.3 (325) | | |
| Stroke | 26.6 (686) | 26.7 (479) | 26.3 (207) | | |
| Asymptomatic | 11.1 (286) | 12.2 (219) | 8.5 (67) | | |
| Ocular | 4.2 (108) | 16.3 (292) | 18.7 (147) | | |
| Retinal infarction | 1.8 (46) | 2.0 (36) | 1.3 (10) | | |
| Other | 17.0 (439) | 4.3 (78) | 3.8 (30) | | |
| Intraplaque haemorrhage (% (n)) | | | | <0.001 | 13.3 |
| Yes | 61.3 (1379) | 64.9 (1019) | 53.1 (360) | | |
| No | 38.7 (870) | 35.1 (552) | 64.9 (1019) | | |
| History of CAD (% (n)) | | | | <0.001 | 0.2 |
| History of CAD | 31.7 (822) | 35.4 (639) | 23.3 (183) | | |
| No history of CAD | 68.3 (1768) | 64.6 (1164) | 76.7 (604) | | |
| History of Stroke (% (n)) | | | | 0.438 | 0.1 |
| History of Stroke | 32.3 (837) | 32.8 (591) | 31.1 (246) | | |
| No history of Stroke | 67.7 (1756) | 67.2 (1212) | 68.9 (544) | | |

**Supplemental Table 2: Performance of each stain in CEA cohort.** Table showing the AUC, accuracy, and F1-score of the test set over 10 k-fold for each stain.

| Stains | Sample Size | Dataset | AUROC (%, std) | Accuracy (%, std) | F1-score (%, std) |
|---|---|---|---|---|---|
| HE | n=1,715<br>IPH/no= 704<br>IPH/yes= 1,011 | train=1,373<br>val=171<br>test=171 | 0.8648 ± 0.0172 | 0.7789 ± 0.0238 | 0.8102 ± 0.0238 |
| FIBRIN | n=628<br>IPH/no= 181<br>IPH/yes= 447 | train=502<br>val=63<br>test=63 | 0.8495 ± 0.0350 | 0.76 ± 0.0580 | 0.7599 ± 0.0482 |
| EVG | n=1,743<br>IPH/no= 713<br>IPH/yes= 1,030 | train=1,394<br>val=174<br>test=175 | 0.8403 ± 0.0305 | 0.7627 ± 0.0312 | 0.7661 ± 0.0288 |
| SR | n=1,422<br>IPH/no= 576<br>IPH/yes= 846 | train=1,136<br>val=143<br>test=143 | 0.8163 ± 0.0232 | 0.7302 ± 0.0343 | 0.7936 ± 0.0248 |
| CD68 | n=1728<br>IPH/no= 705<br>IPH/yes= 1,023 | train=1,384<br>val=172<br>test=172 | 0.7755 ± 0.0311 | 0.7238 ± 0.0299 | 0.7669 ± 0.0210 |
| CD34 | n=1,446<br>IPH/no= 592<br>IPH/yes= 854 | train=1,158<br>val=144<br>test=144 | 0.7722 ± 0.0452 | 0.6962 ± 0.0499 | 0.6867 ± 0.0700 |
| CD66b | n=947<br>IPH/no= 346<br>IPH/yes= 601 | train=757<br>val=95<br>test=95 | 0.7696 ± 0.0620 | 0.7129 ± 0.0569 | 0.7980 ± 0.0386 |
| GLYCC | n=1187<br>IPH/no= 427<br>IPH/yes= 760 | train=949<br>val=119<br>test=119 | 0.7523 ± 0.0662 | 0.685 ± 0.0532 | 0.6703 ± 0.0608 |
| SMA | n=1718<br>IPH/no= 703<br>IPH/yes= 1,015 | train=1,374<br>val=172<br>test=172 | 0.6838 ± 0.0575 | 0.6333 ± 0.602 | 0.7426 ± 0.0297 |

**Supplemental Table 3: Performance of each stain combination.** Table showing the AUC, accuracy, and F1-score of the test set over 10 k-fold for each stain combo in an ensemble logistic regression model.

| Stains | Sample Size | Dataset (k-fold=10) | AUROC (%, std) | Accuracy (%, std) | F1-score (%, std) |
|---|---|---|---|---|---|
| HE + CD68 | n=1,691<br>IPH/no=694<br>IPH/yes=997 | train=1,521<br>test=170 | 0.9149 ± 0.0162 | 0.8368 ± 0.0228 | 0.8625 ± 0.0196 |
| HE + EVG | n=1,715<br>IPH/no=704<br>IPH/yes=1,011 | train=1,543<br>test=172 | 0.9149 ± 0.0171 | 0.8292 ± 0.0242 | 0.8531 ± 0.0256 |
| HE + FIBRIN | n=688<br>IPH/no=197<br>IPH/yes=471 | train=619<br>test=69 | 0.9074 ± 0.0216 | 0.8415 ± 0.0448 | 0.8883 ± 0.0349 |
| HE + CD66b | n=918<br>IPH/no=337<br>IPH/yes=581 | train=826<br>test=92 | 0.8983 ± 0.0307 | 0.8072 ± 0.0495 | 0.8472 ± 0.0393 |
| HE + CD34 | n=1,401<br>IPH/no=578<br>IPH/yes=823 | train=1,260<br>test=141 | 0.8976 ± 0.0293 | 0.8137 ± 0.0320 | 0.8409 ± 0.0371 |
| HE + SR | n=1,399<br>IPH/no=570<br>IPH/yes=829 | train=1,259<br>test=140 | 0.8975 ± 0.0387 | 0.8241 ± 0.0398 | 0.8531 ± 0.0355 |
| HE + SMA | n=1,687<br>IPH/no=693<br>IPH/yes=994 | train=1,518<br>test=169 | 0.8775 ± 0.0215 | 0.7961 ± 0.0272 | 0.8270 ± 0.0265 |
| HE + GLYCC | n=1,289<br>IPH/no=489<br>IPH/yes=800 | train=1,160<br>test=129 | 0.8681 ± 0.0240 | 0.7898 ± 0.0262 | 0.8313 ± 0.0246 |

**Supplemental Table 4: Performance of each stain in the FEA cohort.** Table showing the AUC, accuracy, and F1-score of femoral (FEA) cohort for each stain.

| Stains | Sample Size | AUROC | Accuracy (%) | F1-score |
|---|---|---|---|---|
| EVG | n=766<br>IPH/no=374<br>IPH/yes=392 | 0.83208 | 0.74935 | 0.77251 |
| SR | n=740<br>IPH/no=361<br>IPH/yes=379 | 0.83075 | 0.75135 | 0.76942 |
| HE | n=786<br>IPH/no=375<br>IPH/yes=411 | 0.81333 | 0.73791 | 0.74505 |
| SMA | n=759<br>IPH/no=371<br>IPH/yes=388 | 0.77073 | 0.69697 | 0.71250 |
| FIBRIN | n=177<br>IPH/no=60<br>IPH/yes=117 | 0.75840 | 0.67797 | 0.74208 |
| CD68 | n=705<br>IPH/no=337<br>IPH/yes=368 | 0.75588 | 0.69362 | 0.72308 |
| GLYCC | n=497<br>IPH/no=213<br>IPH/yes=284 | 0.73922 | 0.69416 | 0.73145 |
| CD34 | n=375<br>IPH/no=151<br>IPH/yes=224 | 0.67884 | 0.61600 | 0.64878 |

**Supplemental Table 5: Performance at different thresholds in the HeCES2 cohort.** Table showing the AUC, accuracy, and F1-score of H&E model for HeCES2 cohort at differing threshold. HeCES2 scores model-IPH as percentage of the plaque. The different thresholds of the manually scored IPH area determine when a sample gets an 'IPH yes' label (e.g. a threshold of 2% classifies IPH area > 2% as 'IPH yes', otherwise <= 2% as 'IPH no').

| Threshold (%) | Sample Size | AUROC | Accuracy (%) | F1-score |
|---|---|---|---|---|
| 0.0 | IPH/no=338<br>IPH/yes=138 | 0.78081 | 0.74790 | 0.58621 |
| 1.0 | IPH/no=365<br>IPH/yes=111 | 0.80871 | 0.77101 | 0.58555 |
| 2.0 | IPH/no=374<br>IPH/yes=102 | 0.79666 | 0.76050 | 0.55118 |
| 5.0 | IPH/no=397<br>IPH/yes=79 | 0.80512 | 0.75420 | 0.49351 |
| 10.0 | IPH/no=414<br>IPH/yes=62 | 0.82854 | 0.75630 | 0.45794 |
| 15.0 | IPH/no=425<br>IPH/yes=51 | 0.87672 | 0.75420 | 0.42365 |
| 20.0 | IPH/no=434<br>IPH/yes=42 | 0.89450 | 0.75210 | 0.39175 |
| 25.0 | IPH/no=441<br>IPH/yes=35 | 0.92536 | 0.74580 | 0.35294 |

**Supplemental Table 6: AtherOMICS manual annotation alignment with model-IPH heatmap across varying thresholds.** Table showing the correlation between the manual annotation and the model-IPH heatmap areas, DICE score, sensitivity, specificity.

| Threshold | Area correlation (manual vs model) | DICE | Sensitivity | Specificity |
|---|---|---|---|---|
| 0.50 | 0.811 | 0.2138 ± 0.1712 | 0.3897 ± 0.2612 | 0.7824 ± 0.2322 |
| 0.55 | 0.882 | 0.3667 ± 0.2573 | 0.3703 ± 0.2458 | 0.9753 ± 0.0193 |
| 0.60 | 0.884 | 0.3701 ± 0.2516 | 0.3574 ± 0.2373 | 0.9803 ± 0.0163 |
| 0.65 | 0.885 | 0.3684 ± 0.2464 | 0.3465 ± 0.2296 | 0.9831 ± 0.0147 |
| 0.70 | 0.884 | 0.3657 ± 0.2403 | 0.3368 ± 0.2230 | 0.9851 ± 0.0136 |
| 0.75 | 0.883 | 0.3621 ± 0.2357 | 0.3274 ± 0.2177 | 0.9866 ± 0.0128 |
| 0.80 | 0.881 | 0.3576 ± 0.2303 | 0.3171 ± 0.2129 | 0.9880 ± 0.0120 |
| 0.85 | 0.880 | 0.3523 ± 0.2256 | 0.3055 ± 0.2071 | 0.9891 ± 0.0113 |
| 0.90 | 0.878 | 0.3463 ± 0.2188 | 0.2940 ± 0.2003 | 0.9904 ± 0.0107 |
| 0.95 | 0.873 | 0.3359 ± 0.2104 | 0.2786 ± 0.1930 | 0.9918 ± 0.0099 |

**Supplemental Table 7: AtherOMICS manual versus model annotation metrics at threshold 0.75 across varying IPH area sizes**. Table showing the DICE score, sensitivity, and specificity between the manual annotation and the model-IPH heatmap at increasing model-IPH area sizes.

| Model-IPH area size | DICE | Sensitivity | Specificity |
|---|---|---|---|
| Small (0-5%) | 0.1719 ± 0.1748 | 0.2404 ± 0.2430 | 0.9871 ± 0.0112 |
| Medium (5-25%) | 0.4416 ± 0.1562 | 0.3530 ± 0.1703 | 0.9866 ± 0.0143 |
| Large (25-50%) | 0.5804 ± 0.2127 | 0.4508 ± 0.1906 | 0.9861 ± 0.0128 |
| Very Large (50%+) | 0.6665 ± 0.1457 | 0.5186 ± 0.1679 | 0.9825 ± 0.0108 |

**Supplemental Table 8**: **Correlation between plaque's stained ratio and the model-IPH area size.** Table showing the correlation (r-score) and significance (p-value) per stain.

| Stain | Correlation (r-score) | p-value |
|---|---|---|
| HE (n=1698) | -0.3599 | $4.389\times10^{-53}$ |
| SMA (n=1685) | -0.3028 | $4.534\times10^{-37}$ |
| GLYCC (n=1247) | 0.2646 | $2.017\times10^{-21}$ |
| FIBRIN (n=688) | 0.2969 | $1.801\times10^{-15}$ |
| EVG (n=1430) | -0.1782 | $1.131\times10^{-11}$ |
| SR (n=1396) | -0.0770 | $4.014\times10^{-3}$ |
| CD66b (n=769) | 0.0640 | 0.07725 |
| CD68 (n=1687) | -0.0316 | 0.19454 |
| CD34 (n=1338) | -0.0170 | 0.53558 |

**Supplemental Table 9**: **Differential composition between model-IPH positive plaque samples compared to model-IPH negative plaque samples.** Table showing stain ratio in model-IPH-positive and negative plaque samples, with Mann-Whitney p-value and effect size (r-score).

| Stain | IPH negative (mean, stdev) | IPH positive (mean, stdev) | Mann-Whitney p-value | Effect size r-score |
|---|---|---|---|---|
| SMA | 0.40440 (0.08193) | 0.14235 (0.11665) | $1.689\times10^{-234}$ | -0.786 |
| HE | 0.41922 (0.08375) | 0.31645 (0.10225) | $1.090\times10^{-92}$ | -0.490 |
| GLYCC | 0.06243 (0.05153) | 0.22878 (0.15824) | $4.195\times10^{-91}$ | 0.556 |
| SR | 0.29460 (0.19804) | 0.11388 (0.18570) | $2.217\times10^{-87}$ | -0.522 |
| FIBRIN | 0.05715 (0.05251) | 0.16840 (0.10685) | $5.015\times10^{-77}$ | 0.628 |
| CD34 (LRP) | 0.37692 (0.07669) | 0.26859 (0.12808) | $2.302\times10^{-63}$ | -0.461 |
| EVG | 0.46837 (0.27066) | 0.28834 (0.29112) | $1.867\times10^{-36}$ | -0.326 |
| CD68 | 0.31644 (0.10284) | 0.26022 (0.11697) | $9.928\times10^{-25}$ | -0.247 |
| CD66b | 0.18061 (0.07579) | 0.15261 (0.09557) | $6.389\times10^{-6}$ | -0.154 |

**Supplemental Table 10**: **Imputation metrics.** Missingness ratio and certainty of imputed values for each variable (details on imputation process in Methods).

| **Variable** | **Missingness ratio** | **Certainty of imputed values** |
|---|---|---|
| CRP | 0.532 | 0.7825 |
| HDL | 0.400 | 0.9219 |
| Stenosis contralateral | 0.094 | 0.9835 |
| Symptoms (2 bins) | 0.031 | 0.9625 |
| History of Myocardial Infarction | 0.017 | 0.9054 |
| Anti-coagulant use | 0.007 | 0.8737 |
| Aspirin use | 0.005 | 0.9223 |
| Statin and/or LLD use | 0.004 | 0.9850 |
| Dipyridamole use | 0.003 | 1.0000 |
| Hypertension | 0.001 | 1.0000 |

**Supplemental Table 11**: **Genetic associations of model-IPH-related genes**. Gene name, rsID, Study DOI and corresponding trait of variants whose associated gene (through Locus2Gene) is significantly dysregulated in the model-IPH differential gene expression analysis.

| Gene name | rsID | Study DOI | Trait |
|---|---|---|---|
| ATP2B1 | rs2681492 | doi.org/10.1038/s43587-021-00051-5 | Hypertension |
| CACNB2 | rs12258967 | doi.org/10.1038/s43587-021-00051-5 | Hypertension |
| LPA | rs140570886 | doi.org/10.1038/s43587-021-00051-5 | High cholesterol |
| LSP1 | rs569550 | doi.org/10.1038/s43587-021-00051-5 | Hypertension |
| MMP9 | rs6065906 | doi.org/10.1038/s41588-021-00892-1 | HDL cholesterol levels |
| TRIB1 | rs10555326 | doi.org/10.1038/s43587-021-00051-5 | High cholesterol |

**Supplemental Table 12**: **Clinical Athero-Express characteristic used in the Cox Proportional Hazards Survival Regression model.** Each characteristic is specified by the Athero-Express database variable (for future reference) and additional further explanation

| **Name** | **Database variable** | **Further explanation** |
|---|---|---|
| Age | Age | - |
| Gender | Gender | - |
| Symptoms (2 bins) | Symptoms.Update2G | Symptoms spilt in 'Symptomatic' and 'Asymptomatic' |
| Statin and/or LLD use | Med.Statin.LLD | - |
| History of Myocardial Infarction (MI) | MI_Dx | - |
| Aspirin use | Med.acetylsal | - |
| Anti-coagulant use | Med.anticoagulants | - |
| Dipyridamole use | Med.dipyridamole | - |
| Hypertension | Hypertension.composite | Hypertension and/or using hypertension drugs |
| Stenosis contralateral | stenosis_con_bin | - |
| CRP | CRP_avg | Average CRP value [ug/ml] |
| HDL | HDL_final | HDL-cholesterol [mmol/L] |
| Composite endpoint | epcom.3years | Included composite endpoints within 3 years of follow-up. |
| Time to composite endpoint | ep_com_t_3years | Time to endpoints within 3 years. |
| Intraplaque Hemorrhage (manually scored) | IPH.bin | IPH scored as Yes/No |
| Intraplaque Hemorrhage (predicted by MIL model) | IPH | IPH scored as Yes/No |
| IPH probability (predicted by MIL model) | prob | IPH probability scored between 0 and 1 |
| IPH area (quantified by MIL model) | area | IPH area scored as ratio of total plaque between 0 and 1 |

**Supplemental Table 13: Histological staining protocols and antibodies.** CC1 and CC2 are the buffers of the Ventana staining system used for antigen retrieval. CC1 is an EDTA buffer and CC2 is citrate. In addition, enzyme pretreatment is also available. RTU = Ready To Use. N/a: not applicable.

| STAIN | Stainer | Protocol | Antibody | Dilution | Company | Clone | Clonality |
|---|---|---|---|---|---|---|---|
| CD3 | Ventana (Roche) | 24CC1/32' primary | CD3 | 1/100 | Agilent Dako A0452 | | polyclonal |
| CD34 | Ventana (Roche) | 32'CC1/32' primary | CD34 | RTU | Ventana Q bend 10 | | polyclonal |
| CD66b | Manually | citrate | CD66b | 1/100 | Serotec | | polyclonal |
| CD68 | Ventana (Roche) | 24CC1/32' primary | CD68 | RTU | Roche 790-2931 | | monoclonal |
| GLYCC | Ventana (Roche) | 24CC1/32' primary | glyc.c | 1/800 | Agilent Dako | Ret F40 | |
| SMA | Ventana (Roche) | 24CC1/32' primary | aSMA | 1/20000 | Nordic Biosite BSH-7459-1 | BS66 | |
| EVG | Artisan Link Pro Special Staining System | Kit from Artisan | n/a | n/a | | n/a | n/a |
| Fibrin | Manually | Mallory | n/a | n/a | | n/a | n/a |
| HE | Leica HistoCore SPECTRA ST Stainer | | n/a | n/a | | n/a | n/a |
| SR | Artisan Link Pro Special Staining System | Kit from Artisan | n/a | n/a | | n/a | n/a |

**Supplemental Table 14: Baseline characteristics of the HeCES2 cohort.** Values are averages with standard deviation (SE) unless otherwise noted**.**

| Characteristic | All patients (n=485) | Male (n=330) | Female (n=155) | p-value | Missing (%) |
|---|---|---|---|---|---|
| Age (y (SE)) | 69.7 (0.39) | 69.03 (0.45) | 71.1 (0.72) | 0.012 | 0.0 |
| BMI (kg/m2 (SE)) | 27.6 (0.21) | 27.4 (0.21) | 27.9 (0.46) | 0.301 | 0.0 |
| Regular Smoker (% (n)) | 27.8 (135) | 26.1 (86) | 31.6 (49) | 0.203 | 0.0 |
| Diabetes mellitus (% (n)) | 33.6 (163) | 34.2 (113) | 32.3 (50) | 0.666 | 0.0 |
| Hypertension (% (n)) | 84.9 (412) | 84.5 (279) | 85.8 (133) | 0.717 | 0.0 |
| Hypertensive drugs (% (n)) | 82.5 (400) | 81.8 (270) | 83.9 (130) | 0.579 | 0.0 |
| Anticoagulants (% (n)) | 16.1 (78) | 10.9 (36) | 7.7 (12) | 0.329 | 0.0 |
| Use of statin and/or LLD (% (n)) | 84.8 (384) | 83.4 (262) | 87.8 (122) | 0.483 | 6.6 |
| Symptoms (% (n)) | | | | 0.131 | 0.0 |
| Yes | 65.6 (318) | 63.3 (209) | 70.3 (109) | | |
| No | 34.4 (167) | 36.7 (121) | 29.7 (46) | | |
| Intraplaque haemorrhage (% (n)) | | | | <0.001 | 1.2 |
| Yes | 29.0 (139) | 34.5 (112) | 17.5 (27) | | |
| No | 71.0 (346) | 65.5 (218) | 82.5 (128) | | |
| History of MI, CAD or coronary intervention (% (n)) | 37.3 (181) | 43.9 (145) | 23.2 (36) | <0.001 | 0.0 |
| History of Stroke or TIA (% (n)) | 43.2 (209) | 46.5 (153) | 36.1 (56) | 0.065 | 0.0 |

**Supplemental Table 15: Baseline characteristics of the AtherOMICS cohort.** Values are averages with standard deviation (SE) unless otherwise noted.

| Characteristic | All patients (n=98) | Male (n=68) | Female (n=30) | p-value | Missing (%) |
|---|---|---|---|---|---|
| Age (y (SE)) | 72.09 (8.80) | 72.56 (9.02) | 71.03 (8.32) | 0.432 | 0.0 |
| BMI (kg/m2 (SE)) | 26.154 (4.401) | 26.277 (3.336) | 25.865 (6.296) | 0.675 | 1.0 |
| Diabetes mellitus (% (n)) | 30.6 (30) | 23.5 (16) | 46.7 (14) | 0.055 | 0.0 |
| Smoking status (% (n)) | | | | 0.263 | 0.0 |
| Yes, current | 26.5 (26) | 26.5 (18) | 26.7 (8) | | |
| Yes, former | 34.7 (34) | 32.4 (22) | 40.0 (12) | | |
| No | 30.6 (30) | 29.4 (20) | 33.3 (10) | | |
| Unknown | 8.2 (8) | 11.8 (8) | 0.0 (0) | | |
| Symptoms (% (n)) | | | | 0.610 | 0.0 |
| Yes | 61.2 (60) | 58.8 (40) | 66.7 (20) | | |
| No | 38.8 (38) | 41.2 (28) | 33.3 (10) | | |

**Supplemental Table 16**: Long-range LD blocks excluded during GWAS quality control step (genome b38).

**(SEE SEPARATE EXCEL FILE)**

**Supplemental Table 17:** Baseline characteristics of 8 spatial samples from the Athero-Express Biobank Study. Values are averages with standard deviation (SE) unless otherwise noted.

| Characteristic | All patients (n=8) | Missing (%) |
|---|---|---|
| Age (y (SE)) | 71.88 (8.87) | 0.0 |
| Sex - male (% (n)) | 50.0 (4) | 0.0 |
| BMI (kg/m2 (SE)) | 25.372 (4.406) | 12.5 |
| Current Smoker (% (n)) | 37.5 (3) | 0.0 |
| Diabetes mellitus (% (n)) | 25.0 (2) | 0.0 |
| Hypertension (% (n)) | 100.0 (8) | 0.0 |
| Hypertensive drugs (% (n)) | 87.5 (7) | 0.0 |
| Anticoagulants (% (n)) | 0 (0) | 0.0 |
| Use of statin and/or LLD (% (n)) | 75.0 (6) | 0.0 |
| Glomerular filtration rate (mL/min (SE)) | 87.62 (42.27) | 37.5 |
| Restenosis (% (n)) | 0 (0) | 0.0 |
| Symptoms (% (n)) | | 0.0 |
| TIA | 12.5 (1) | |
| Stroke | 37.5 (3) | |
| Asymptomatic | 25.0 (2) | |
| Ocular | 12.5 (1) | |
| Retinal infarction | 12.5 (1) | |
| Other | 0.0 (0) | |
| History of CAD (% (n)) | | 0.0 |
| History of CAD | 0.0 (0) | |
| No history of CAD | 100.0 (8) | |
| Manual IPH binary - Yes (% (n)) | 50.0 (4) | 0.0 |
| Model IPH binary - Yes (% (n)) | 75.0 (6) | 0.0 |
| Model IPH probability (% (n)) | 0.575 (0.215) | 0.0 |
| Model IPH area ratio (% (n)) | 0.100 (0.062) | 0.0 |

**Supplemental Table 18**: Library preparation and sequencing metrics for the 8 Visium HD samples.

| Sample Pair | Visium Slide | Position | Library Tag | Fragment Size (bp) | Library Conc. (nM) | Index Name | Index i7 (5'→3') | Flowcell_Lane | Raw Reads | Raw Data (Gb) | Error (%) | Q20 (%) | Q30 (%) | GC (%) | Bins Detected (tissue-covered) |
|---|---|---|---|---|---|---|---|---|---|---|---|---|---|---|---|
| Pair-1 | H1-V9C329D | A1 | 1A | 320 | 47.53 | SI-TS-B1 | TATGACGAGT | 22GKHGLT4_L8 | 564,843,180 | 84.73 | 0.06 | 76.97 | 60.99 | 45.54 | 378,466 |
| Pair-1 | H1-V9C329D | D1 | 1B | 331 | 80.36 | SI-TS-B2 | ACCTGGTACA | 22GKHGLT4_L8 | 570,017,532 | 85.50 | 0.06 | 77.20 | 61.13 | 45.90 | 306,339 |
| Pair-2 | H1-MTNK24G | A1 | 2A | 325 | 64.54 | SI-TS-B3 | GTCACGGGTG | 22GKHGLT4_L8 | 572,307,680 | 85.85 | 0.06 | 77.77 | 62.03 | 46.62 | 331,024 |
| Pair-2 | H1-MTNK24G | D1 | 2B | 314 | 51.22 | SI-TS-B4 | ACCCGCTCGA | 22GKHGLT4_L8 | 525,646,222 | 78.85 | 0.06 | 76.91 | 60.84 | 45.74 | 288,897 |
| Pair-3 | H1-KKJ4NK9 | A1 | 3A | 328 | 56.33 | SI-TS-B5 | GTGTCTAACT | 22GKHGLT4_L8 | 503,586,614 | 75.54 | 0.06 | 76.91 | 61.11 | 45.27 | 548,906 |
| Pair-3 | H1-KKJ4NK9 | D1 | 3B | 321 | 78.62 | SI-TS-B6 | CGGCCATAGG | 22GKHGLT4_L8 | 574,052,132 | 86.11 | 0.06 | 77.05 | 60.94 | 45.79 | 394,684 |
| Pair-4 | H1-T2788B4 | A1 | 4A | 313 | 78.98 | SI-TS-B7 | CATGCTGCTC | 22GKHGLT4_L8 | 489,279,638 | 73.39 | 0.06 | 77.11 | 61.16 | 45.83 | 476,160 |
| Pair-4 | H1-T2788B4 | D1 | 4B | 341 | 234.84 | SI-TS-A9 | CCACTTACGT | 22GKHGLT4_L8 | 527,169,160 | 79.08 | 0.06 | 76.89 | 61.07 | 45.80 | 354,840 |

Fragment size, library concentration, and index assignments from the sequencing submission tracking sheet. Raw reads, raw data, error rate, Q20/Q30, and GC content are actual sequencing output from the Novogene QC report (flowcell lane 22GKHGLT4_L8, 150 bp single-end reads). Bins detected = number of 2×2 μm bins with tissue coverage, out of 702,244 bins per capture area maximum - the tissues are not completely covering the Visium HD slide. Library Tag to Novogene QC linkage was inferred by matching row order between the tracking sheet and the Novogene report.
